# Neurofunctional correlates of Working Memory across Psychosis Stages: A Systematic Review and Meta-analysis

**DOI:** 10.64898/2026.01.12.26343386

**Authors:** Maria-Mihaela Avram, Lesley Bayly-Bureau, Nicholas R Livingston, Paolo Fusar-Poli, Matthew J Kempton, Joaquim Radua, Mitul Mehta, Gemma Modinos

## Abstract

Working memory (WM) impairments have been reported in different stages of psychosis but whether their neural correlates are shared or stage-specific is unknown. This meta-analysis examined WM-related brain activity across psychosis stages: familial and clinical high-risk for psychosis (at-risk stage), first-episode psychosis (early psychosis stage), and chronic schizophrenia (chronic psychosis stage). PubMed, Ovid, and Web of Science were searched up to July 2025 for functional magnetic resonance imaging (fMRI) studies comparing individuals in each stage and healthy controls during WM. Seed-based d-mapping assessed WM-related fMRI correlates at each stage. Significance was set at family-wise error-corrected p<.05. Forty-two studies were included: 7 in the at-risk stage, 5 in the early psychosis stage, and 30 in the chronic psychosis stage. In chronic psychosis, higher activation relative to controls was observed in the medial prefrontal cortex, rostral anterior cingulate, right insula and superior temporal gyrus, posterior cingulate cortex, left superior temporal and supramarginal gyri. Lower activation in chronic psychosis vs controls was found in the cerebellum, bilateral precuneus, middle temporal gyrus, and thalamus. The early psychosis stage was characterised by lower activation compared to controls in the dorsal anterior cingulate, bilateral caudate nuclei, and inferior frontal gyrus. No significant clusters emerged in the at-risk stage, or across stages. In combined early and chronic psychosis analyses, anterior cingulate cortex activation was positively associated with both antipsychotic dose and illness duration. These findings indicate that disruptions in WM circuitry may evolve after illness onset and may represent a potential biomarker of psychosis staging.

**Highlights:**

- This study revealed distinct brain activity patterns in early and chronic psychosis stages.
- Alterations in brain activity were more widespread in the chronic stage of psychosis.
- Antipsychotic dose and illness duration predicted anterior cingulate cortex activation.
- Distinct neural correlates of working memory may reflect illness progression.

## 1. Introduction

Working memory (WM) impairment is a core cognitive deficit in psychotic disorders such as schizophrenia and has been associated with poor functioning and quality of life (Halverson et al., 2019; Kadakia et al., 2022). WM abnormalities, among other cognitive deficits, have been documented across at-risk and clinical stages of psychosis, from familial high-risk (Bora et al., 2014; Snitz et al., 2006) (FHR, first-degree relatives of individuals with psychosis), clinical high-risk for psychosis(CHR-P) (Catalan et al., 2021), to first-episode psychosis (FEP) (Catalan et al., 2024; Lee et al., 2024) and chronic schizophrenia (Forbes et al., 2009). Furthermore, lower WM performance was found to be associated with subsequent transition to psychosis in individuals at CHR-P (Fusar-Poli et al., 2012; Seidman et al., 2016), and relapse in those with FEP (Rund et al., 2007), which was also associated with worsening of WM performance (Hui et al., 2016; Tao et al., 2023). Overall, these findings suggest that WM deficits may be an early marker of psychosis and warrant investigation into their neural substrate, to understand whether they are state- or trait-related.

Previous meta-analyses reported altered WM-related activation in frontoparietal cortical regions, including middle frontal gyrus and posterior parietal cortex, across different stages of psychosis (at-risk, first-episode, chronic psychosis) (Dutt et al., 2015; Glahn et al., 2005; Wang et al., 2021; Wu and Jiang, 2020; Yao et al., 2024; Zachary Adam Yaple et al., 2021; Zhang et al., 2016). However, results across these meta-analyses are mixed, with substantial inconsistency across studies and stages examined. This may be linked to between-study heterogeneity, such as differing approaches in participant sampling, imaging parameters and scanner type, WM paradigms, and choice of analysis. In addition, differences in meta-analytic methodologies, such as inclusion criteria, number of included studies, and significance thresholding may have had a role. Importantly, all the published neuroimaging meta-analyses on WM in psychosis stages used coordinate-based meta-analytic approaches (Albajes-Eizagirre and Radua, 2018), which, despite enabling the quantitative synthesis of a large body of literature, rely exclusively on reported peaks of activation and sample sizes, thus overlooking considerable amounts of information that does not reach the significance threshold^24^. In contrast, image-based meta-analyses show higher power and sensitivity compared to approaches relying on peak coordinates (Albajes-Eizagirre and Radua, 2018; Radua and Mataix-Cols, 2012; Salimi-Khorshidi et al., 2009), but are limited by the availability of statistical maps from individual studies (Radua and Mataix-Cols, 2012; Salimi-Khorshidi et al., 2009).

Another source of heterogeneity is the inclusion of a wide range of tasks and contrasts, some of which involve WM processing at different phases, such as encoding, maintenance, retrieval (Bittner et al., 2015) or effects of emotional stimuli on WM performance (Becerril and Barch, 2011), which may show differing activity in the brain. Importantly, many studies use contrasts that compare a WM condition with a fixation cross or rest, which may reflect non-WM signals, such as vigilance, motor response or visual processing, potentially overriding the WM effects when included consistently. In addition, previous meta-analyses often include multiple contrasts from the same participants and tasks, which are not accounted for in the analyses (Yao et al., 2024; Zhang et al., 2016).

The present study is an updated fMRI meta-analysis on neural correlates of WM across clinical stages of psychosis. Specifically, we aimed to address the limitations associated with coordinate-based approached by combining peak coordinate data and statistical maps obtained from study authors using seed-based d-mapping (SDM) (Radua et al., 2012b). SDM generates effect size maps accounting for increases and decreases in brain activation and allows the combination of peak coordinates and statistical maps (Albajes-Eizagirre et al., 2019; Radua et al., 2012b), showing a 20% increase in sensitivity of results when at least 3 maps are included (Radua and Mataix-Cols, 2012). To further minimise heterogeneity, we only included studies using one category of WM tasks, namely the ‘n-back’ paradigm. This WM paradigm involves the use of several cognitive processes, such as encoding, maintenance and continuous updating in order to detect a stimulus shown *n* positions previously (Callicott et al., 1998). The n-back task is used frequently in studies exploring groups across psychosis stages (Dutt et al., 2015; Glahn et al., 2005), it has high reproducibility in the neuroimaging literature (Gradin et al., 2010; Mencarelli et al., 2019; Owen et al., 2005) and unified WM task design.

## 2. Methods

This systematic review with meta-analysis was preregistered on PROSPERO (available from: https://www.crd.york.ac.uk/PROSPERO/view/CRD42024538011) and reporting follows the PRISMA guidelines (Table S1).

### 2.1. Searches and study selection

A systematic search was conducted on PubMed, OVID and Web of Science databases for all studies published until July 11^th^ 2025, using the search terms as shown in the Supplement. In addition, the bibliographies of 10 meta-analyses were searched (Dutt et al., 2015; Fusar-Poli, 2012; McTeague et al., 2017; Picó-Pérez et al., 2022; Soldevila-Matías et al., 2022; Wang et al., 2021; Wu and Jiang, 2020; Yao et al., 2024; Z. A. Yaple et al., 2021; Zhang et al., 2016).

The screening of abstracts and full texts was conducted by 2 investigators (MMA and LBB). Studies were included if they explored case-control differences in fMRI activation during the n-back task in the at-risk and clinical stages in psychosis. The at-risk stage comprised individuals who had a first-degree relative with schizophrenia or schizoaffective disorder (FHR) or individuals at CHR-P who met criteria for CAARMS (Yung et al., 2005) (Comprehensive Assessment of At-Risk Mental States) or SIPS (Miller et al., 2002) (Structured interview for Prodromal syndromes). Early and chronic psychosis were defined as individuals who had an established diagnosis of schizophrenia or schizoaffective disorder, based on criteria defined by DSM-IV (Diagnostic and Statistical Manual of Mental Disorders, 4^th^ edition), DSM-V (Schizophrenia Spectrum Disorders), or ICD-10 (International Classification of Diseases, 10th Revision, F20 or F25).

Exclusions at the abstract screening stage were based on the following criteria: (1) there were no mentions/inferences of task-based fMRI in the abstract, (2) there were no mentions/inferences of any population of interest in the abstract, (3) there was an explicit statement of the fMRI task not assessing mainly working memory, (4) there were no mentions/inferences of healthy control (HC) groups, (5) the reported sample size per comparison group was lower than 7 participants per comparison group (Tahmasian et al., 2019), and (6) the abstract was for a systematic review, meta-analysis, conference report, case report/series, preprint, or dissertation.

At the full-text screening stage, studies were excluded if whole-brain results were not reported, the task design included distractors or emotional stimuli, results reflected effects of interventions with no baseline data, data was not normalised in the Montreal Neuroimaging Institute (MNI)(Evans et al., 1993) or Talairach reference spaces (Talairach and Tournoux, 1988). Studies were only included if these reported findings from a contrast that isolated WM effects, such as “2-back>0-back”, “2-back>1-back”, or a group-by-load linear model analysis. Authors of all eligible studies were contacted to request whole-brain statistical maps of the contrasts reported in the papers.

Originally, studies using all WM tasks were considered for the meta-analysis. After screening, most of the eligible studies had used n-back tasks, and thus we decided to only include studies using versions of the n-back task in the meta-analysis, to reduce heterogeneity caused by differences in WM paradigms. Furthermore, a few studies used WM tasks that explored multiple phases of WM. Since using multiple contrasts would bias the findings towards a specific study, we decided to exclude those studies.

#### 2.1.1. Studies with overlapping cohorts

To maximise the independence of data, we employed extra steps to identify studies with overlapping cohorts. First, we suspected overlap when studies exploring a similar group were published by either the same authors or authors from the same institutions. Then, we searched for potential mentions of a previous study based on the same cohort in the methods section. When this information was not stated, we checked for similarities in participant demographics, including age, sex distribution, antipsychotic use, IQ, etc. In one case, we identified 3 studies with similar authors, which were revealed to use data from overlapping cohorts after communicating with the corresponding author.

When studies reporting on overlapping cohorts were eligible, the inclusion was decided based on the following order of priorities: (1) the authors of the study shared a statistical map; (2) the study reported corrected findings, such as using false-discovery rate or family-wise error corrections; (3) the study used a higher sample size; (4) the study had a lower risk of bias.

Further information on screening and data extraction is described in the Supplement.

### 2.2. Quality assessment

The quality assessment was performed using a version of the Newcastle-Ottawa Scale adapted for neuroimaging studies (Gentili et al., 2019). This scale evaluates the reports based on criteria such as study population selection, matching, behavioural manipulation, p-value thresholding and applied corrections. Based on the score, the studies were assigned to low, moderate, or high risk of bias categories.

### 2.3. Meta-analyses

The meta-analyses were conducted in SDM with Permutation of Subject Images (SDM-PSI), version *6.23* (Albajes-Eizagirre et al., 2019) for 3 groups of interest that represented at-risk and clinical stages of psychosis. These comprised (1) at-risk group – studies where participants had FHR or CHR-P, (2) early psychosis group – studies where participants had either FEP or a mean duration of illness (+ SD) lower than 5 years and (3) chronic psychosis group – studies where participants had a diagnosis of schizophrenia or schizoaffective disorder (illness duration mean (years) + SD > 5). Illness duration cut-offs were chosen based on established definitions of early psychosis (Newton et al., 2018) and incorporated standard deviations to minimise misclassification.

The statistical maps obtained from study authors were converted to *T*-maps in SDM where applicable. For the remaining studies, reported whole-brain peak coordinates were used, in addition to reported *T*-values. Where *T*-values were missing, the *Z*- or *p*-values were converted using the online conversion tool (www.sdmproject.com). Studies that reported non-significant differences in activation, and for which statistical maps were not available were included as null findings, following the SDM protocol.

Studies were pre-processed using the SDM built-in command that involved a grey matter mask and the default settings for the fMRI modality (anisotropy =1, voxel size = 2mm, isotropic full-width half maximum = 20) (Pollard et al., 2023). Pre-processing was followed by mean analyses (meta-analyses) using 50 imputations, and family-wise error (FWE) corrections via threshold free cluster enhancement using 1000 permutations, which were part of the default SDM pre-sets. Statistical significance was set at *pFWE* < .05.

All primary meta-analyses included age as a covariate, due to considerable between-study variability in mean ages of participants (see Results), and previous findings of significant effects of age on WM-related fMRI activity (Wang et al., 2019). The age variable was calculated as the weighted mean in cases and control ages. Due to the nature of the method, only bidirectional effects could be inferred. For the purpose of clarity, we classified all results for cases > controls as hyperactivations and for controls > cases as hypoactivations.

#### Secondary analyses

Conjunction and disconjunction analyses, using the “Multimodal” function in SDM, were conducted to examine overlapping regions of activity across psychosis stages. We conducted meta-comparisons for early psychosis > at risk, and chronic psychosis > early psychosis contrasts using the “Linear Model” function. This was done by adding another column to the dataset where one group was allocated 1 and the other 0, simulating the meta-comparison functions from previous versions of SDM (Radua et al., 2010). A threshold of *p_uncorr_* <.0005 was chosen to determine statistical significance, as previously suggested for linear model analyses (Radua et al., 2012b).

Meta-regressions were conducted in each group, also using the “Linear Model” function to explore the effects sex distribution in cases vs controls on brain activation. This was done due to previous evidence on sex differences in fMRI activation during the n-back task in schizophrenia (Elsabagh et al., 2009). In addition, meta-regressions were conducted in the combined early and chronic psychosis group to explore the effects of antipsychotic dose (mean CPZ equivalents) and mean duration of illness (years) on WM-related brain activity. The significance was set at *p < .0005* uncorrected, as suggested previously (Radua et al., 2012a). The calculation of each predictor in meta-regressions is described in the Supplement.

Finally, sensitivity analyses were conducted in each group by excluding the age covariate. Subgroup meta-analyses were conducted for FHR and CHR-P separately. Heterogeneity and publication bias were assessed using the “Extract” and “Bias Test” functions in SDM-PSI, which compute *I^2^* statistics and perform the meta-bias tests, respectively. Robustness of the findings was evaluated using jack-knife sensitivity analyses, iteratively re-running the 3 main meta-analyses by excluding each study once. The significance thresholds of all sensitivity analyses were set as in the main meta-analyses. All the results are presented in the MNI template space.

All descriptive statistics and additional calculations were performed using R Studio, version *4.4.3*. The reports of medians and ranges were transformed into means and standard deviations using the *estmeansd* package, which enables quantile estimation without assuming normal distributions (McGrath et al., 2020). Several studies reported doses for separate antipsychotics, and the number of participants who were prescribed these antipsychotics, thus the *chlorpromazineR* package was used to convert each reported antipsychotic and its dose to CPZ equivalents, based on an international consensus on antipsychotic dosing (Gardner et al., 2010). Following this, a weighted mean of CPZ equivalents was estimated. To account for the participants who did not take medication, the dose variable was again weighted by the proportion of medicated subjects. The same weighting procedure was applied for studies that reported mean CPZ equivalents for medicated participants.

## 3. Results

### 3.1. Included studies

The identification, screening and inclusion of studies is shown in the PRISMA flow diagram, Figure 1 (Page et al., 2021). A total of 42 studies comprising 1270 participants across at-risk and clinical stages in psychosis were included. Authors of 71 studies were initially contacted to request whole-brain statistical maps, of which, authors of 45 studies responded and authors of 13 studies provided 15 statistical maps in total. Four maps from 2 studies were then excluded due to explicit masking of results. As a result, the meta-analysis consisted of 45 datasets derived from the included studies, of which 32 datasets comprised whole-brain peak coordinates, 11 were statistical maps, and 2 were null datasets (non-significant findings and no maps). Detailed characteristics for all included studies are described in Tables 1-3, and summary characteristics are shown in Table 4.

**Figure 1.**
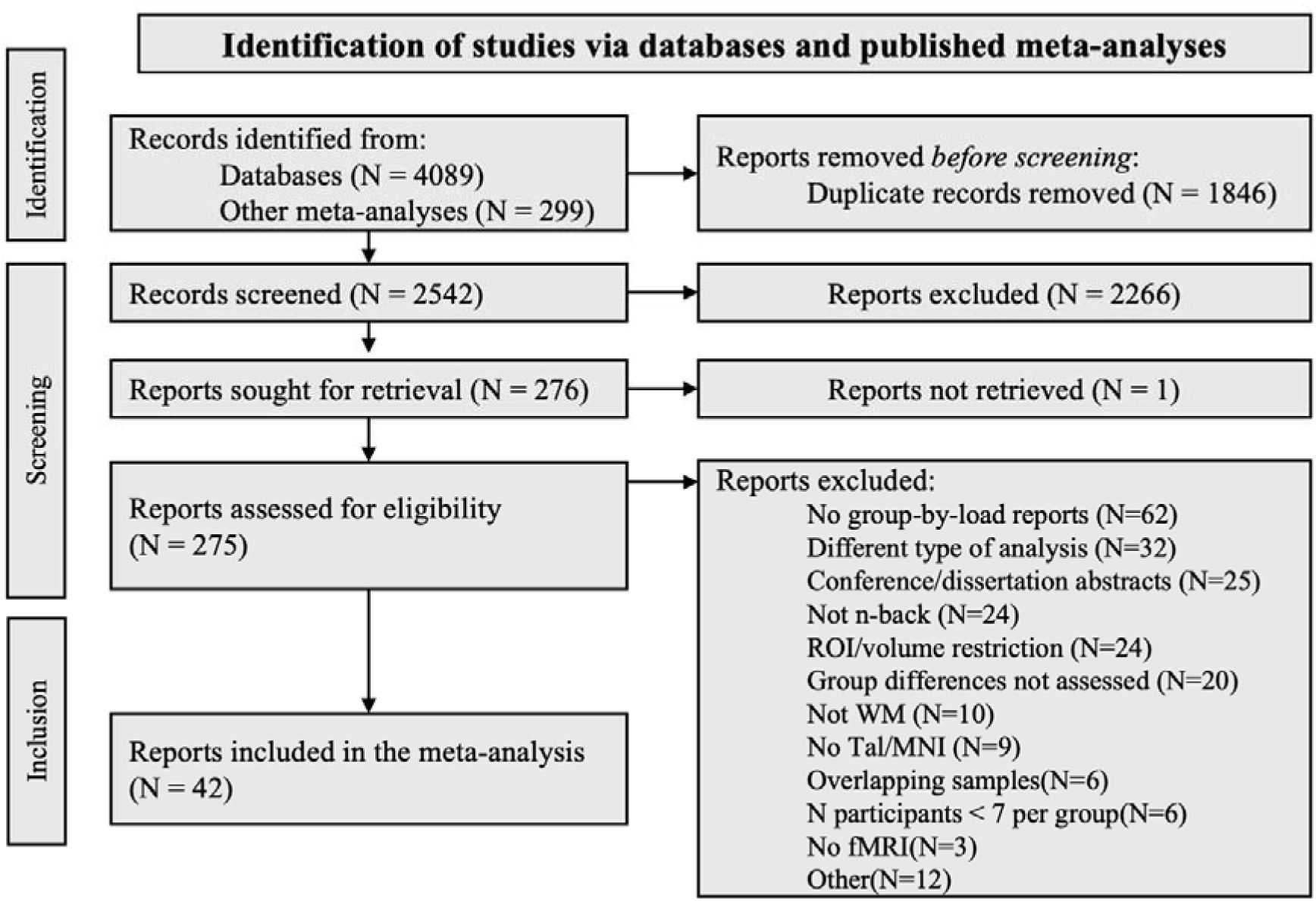
PRISMA flow diagram depicting the study selection process for the meta-analysis.

**Table 1.**
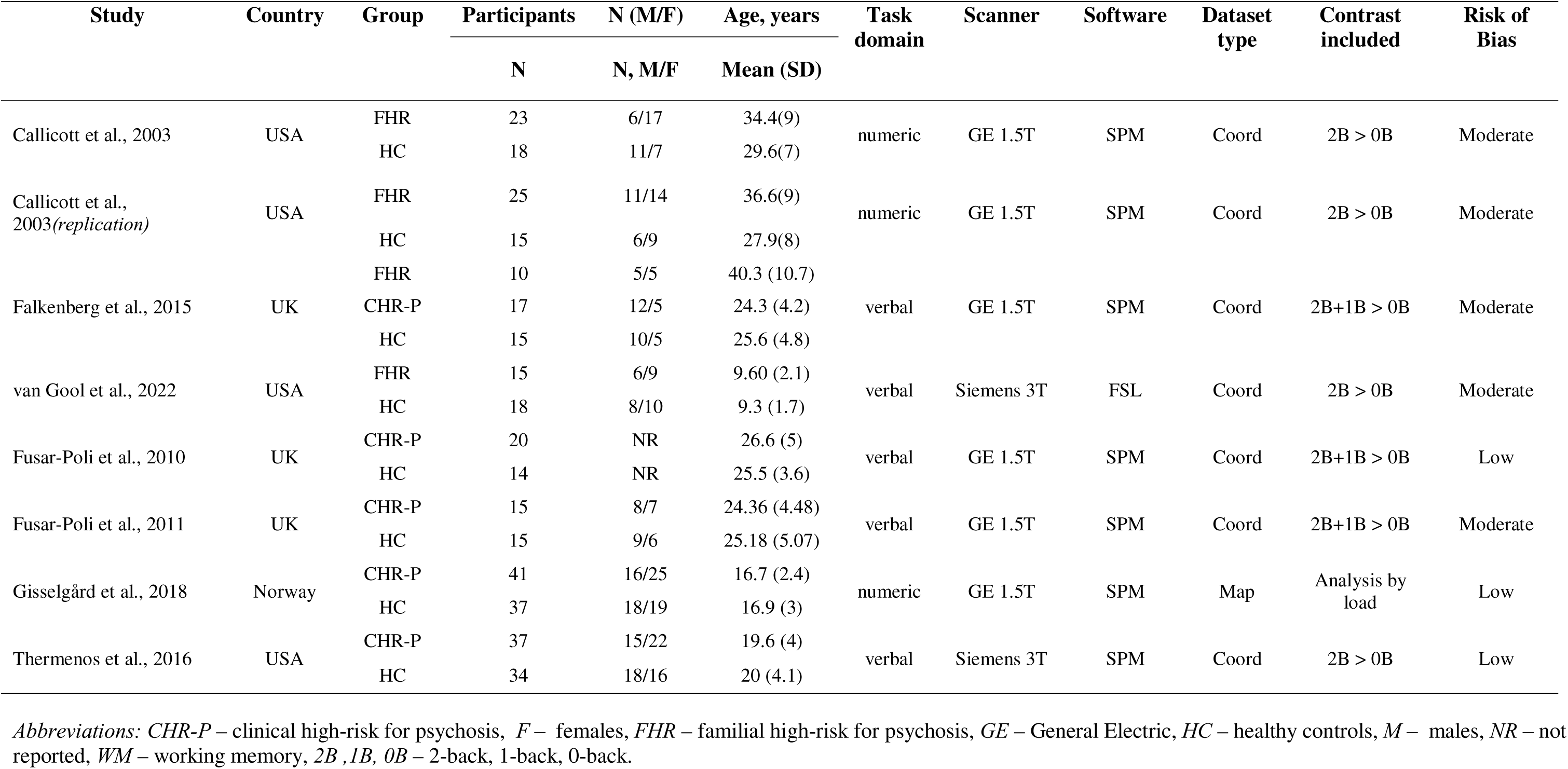
Characteristics of included studies in the at-risk stage.

**Table 2.**
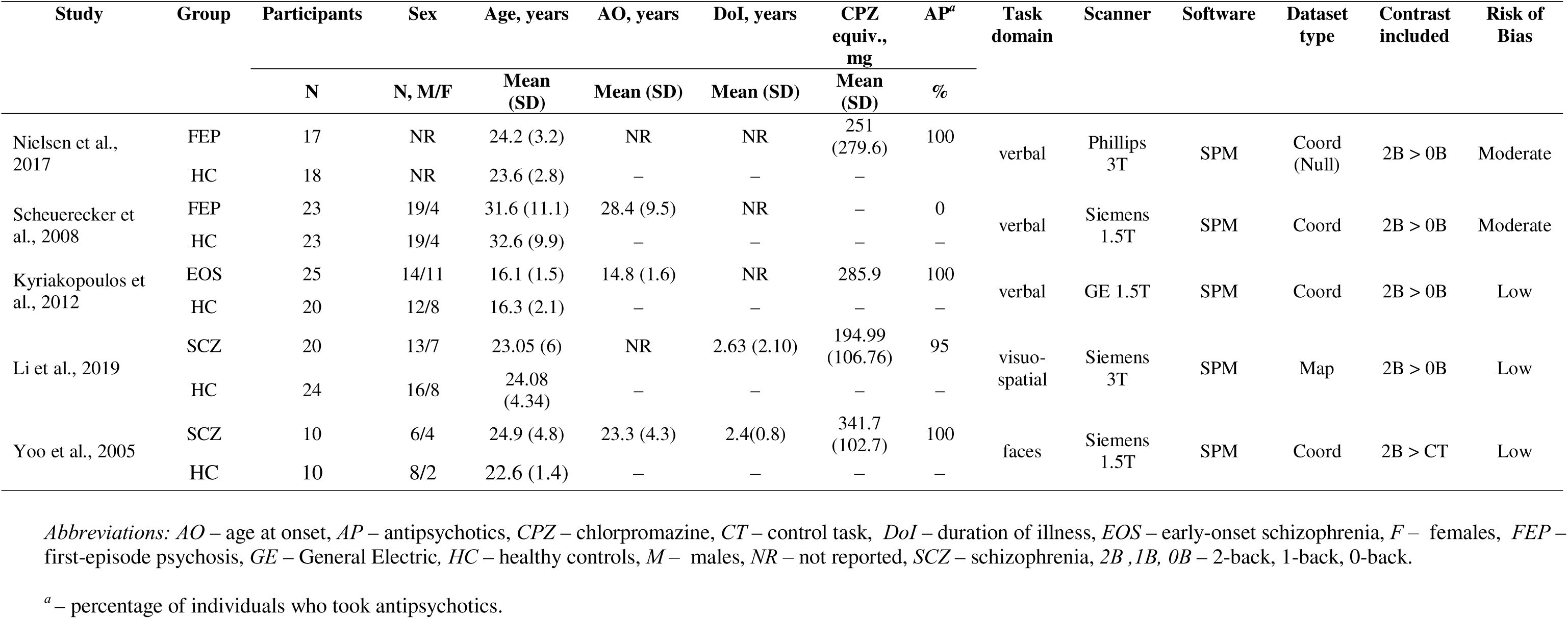
Characteristics of included studies in the early psychosis stage.

**Table 3.**
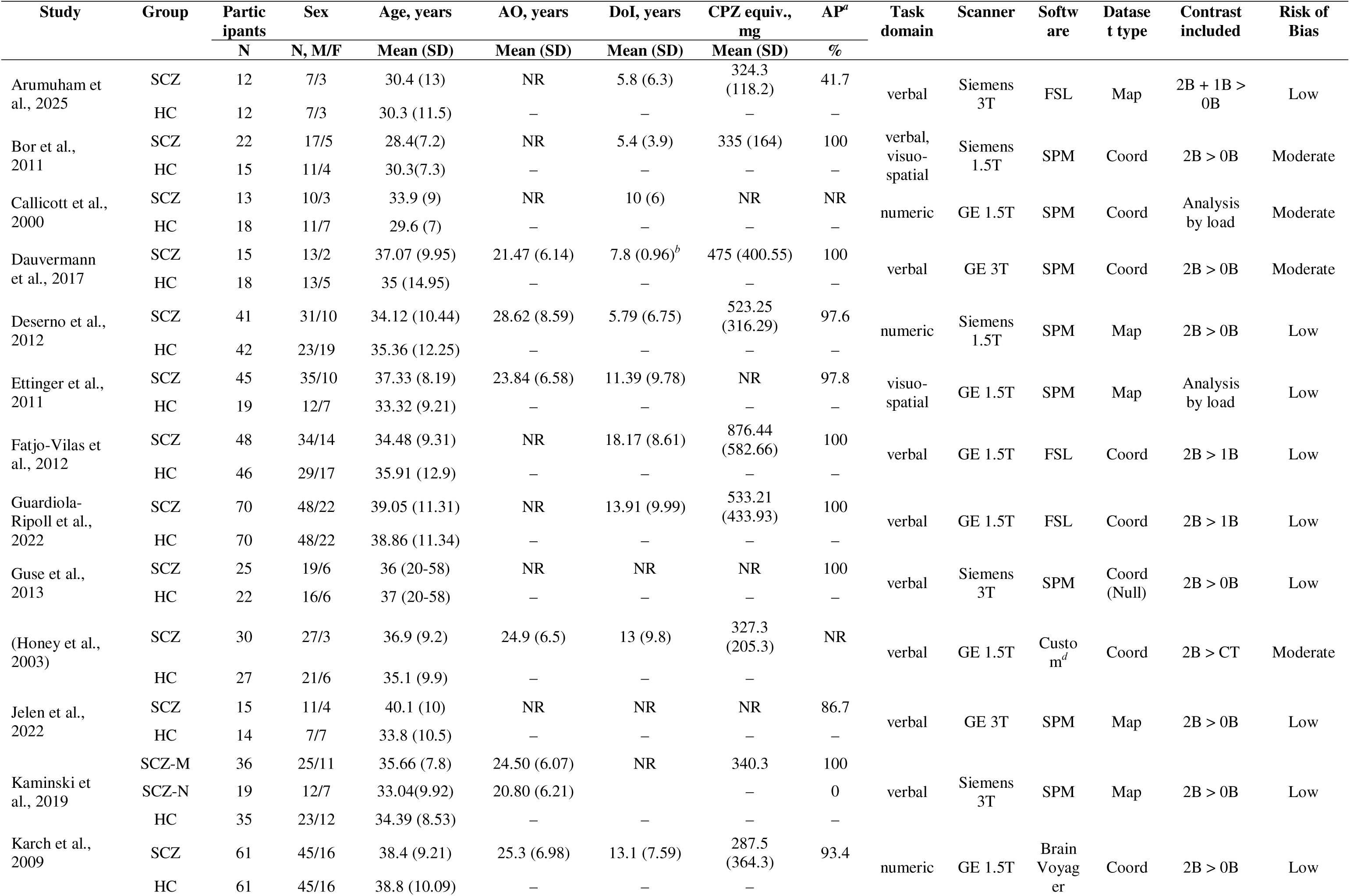
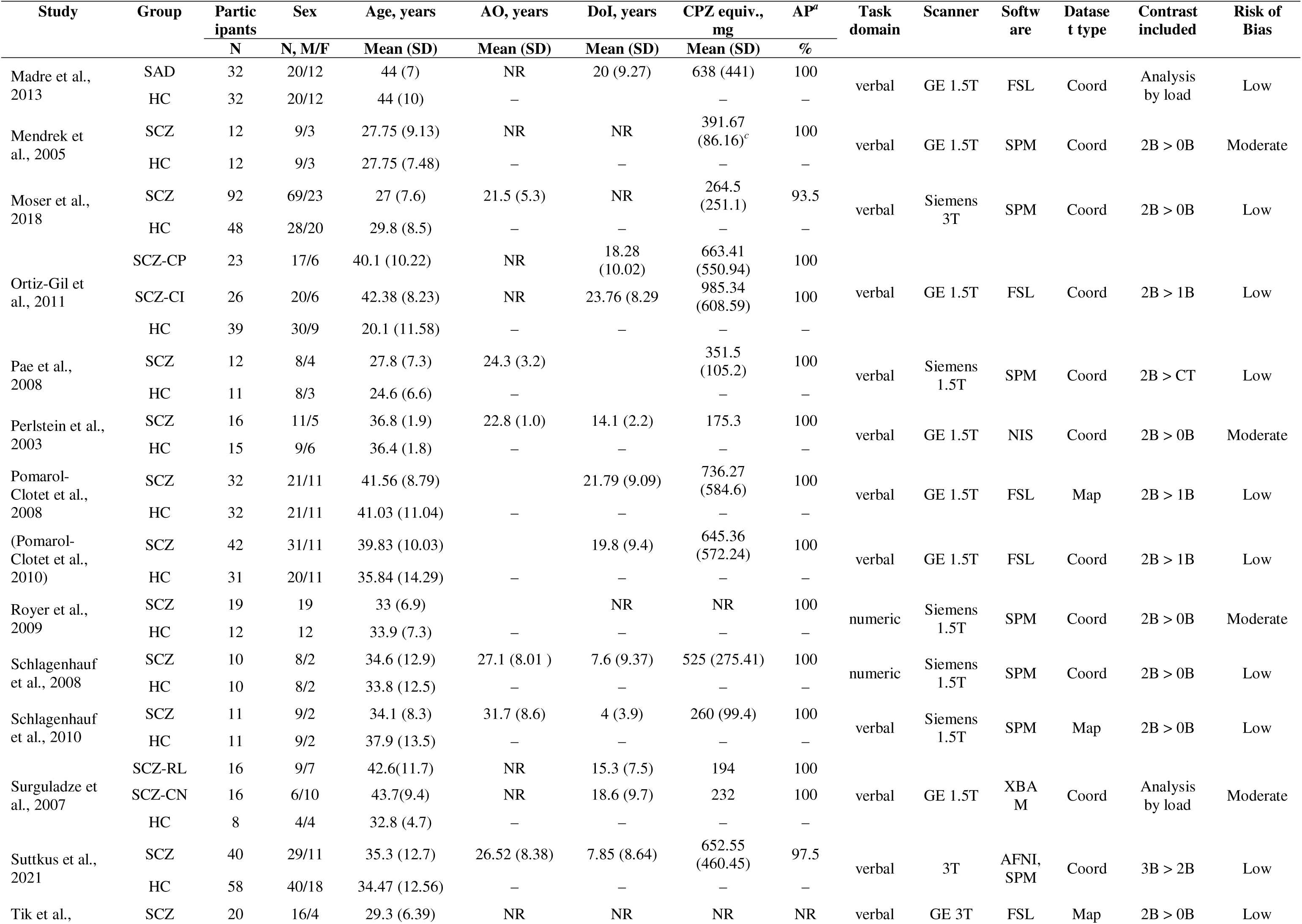
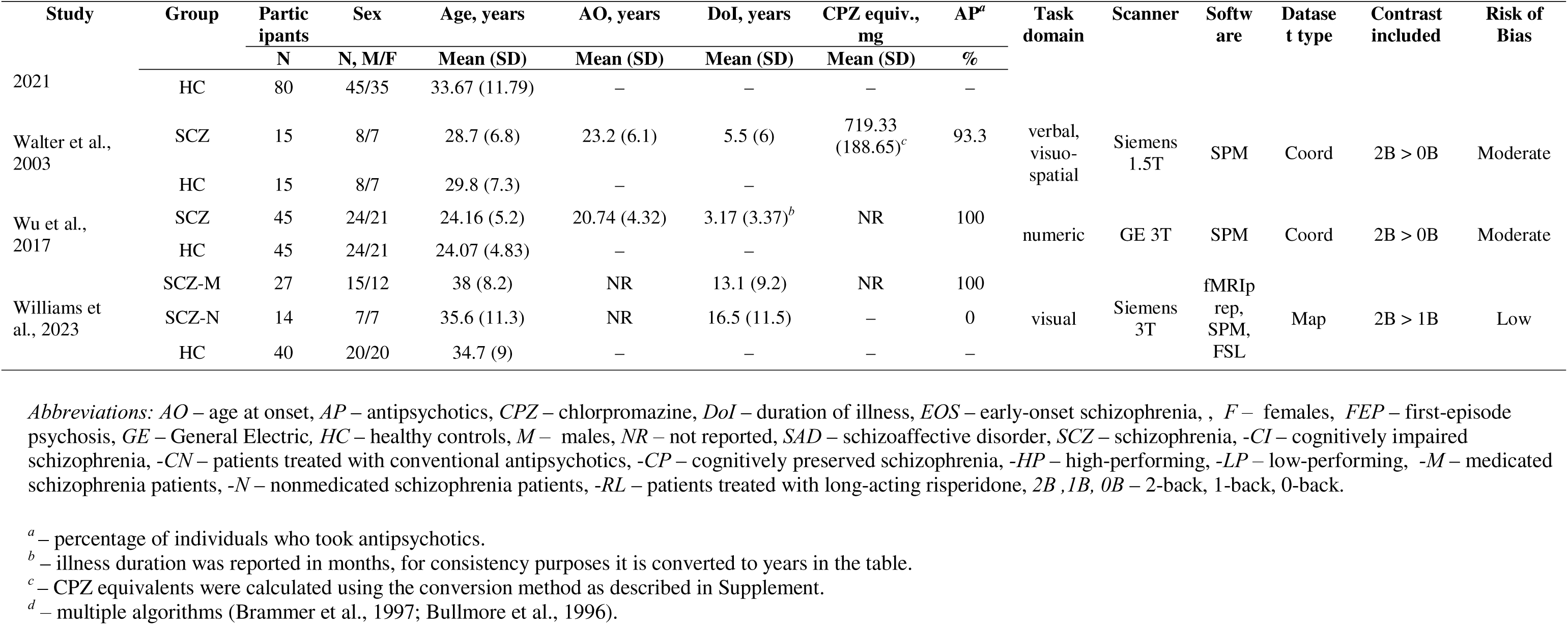
Characteristics of included studies in the chronic psychosis stage.

**Table 4.** Summary statistics of the included studies.

| **Variable** | **At-risk for psychosis**<br>**(7 studies)** |  | **Early psychosis**<br>**(5 studies)** |  | **Chronic psychosis**<br>**(30 studies)** |  |
| --- | --- | --- | --- | --- | --- | --- |
|  | **Cases** | **Controls** | **Cases** | **Controls** | **Cases** | **Controls** |
| **Participants, n** | 203 | 166 | 95 | 95 | 972 | 898 |
| **Age range<sup>a</sup>, years** | 9.6 – 40.3 | 9.3 – 29.6 | 16.1 – 31.6 | 16.3 – 32.6 | 24.16 – 44.0 | 20.4 – 44.0 |
| **Sex<sup>b</sup> (% male)** | 40.91 | 52.63 | 66.67 | 71.43 | 70.83 | 64.44 |

*Abbreviations: HC – healthy controls, NS – non-significant datasets that were included as empty .txt files in the analysis.*
<sup>a</sup> – The values shown represent mean ages from the included studies
<sup>b</sup> – One study from each group did not report sex distribution, these studies were not included in the percentage calculations. More detailed study characteristics are reported in eTables 2,3,4.

### 3.2. Meta-analytic findings

In the meta-analysis for the at-risk stage, no significant WM-related fMRI alterations survived correction. At a less stringent threshold employed in a previous study (*p_uncorr_*<.005; cluster extent=100 voxels; (Yao et al., 2024)), we found a hyperactivation of right middle frontal gyrus in the at-risk individuals compared to controls (x=38, y=52, z=0; *p_uncorr_*=.00005; *pFWE* = .29, Z = 3.9, Figure S1). No findings in the FHR and CHR subgroups separately survived correction (Table S2).

At the early psychosis stage, five significant clusters of hypoactivation relative to controls were identified, in the medial superior frontal gyrus and dorsal ACC, bilateral caudate nuclei and anterior thalamus, and inferior frontal gyri (Table 5; Figure 2A). No clusters showed hyperactivation relative to controls.

**Figure 2.**
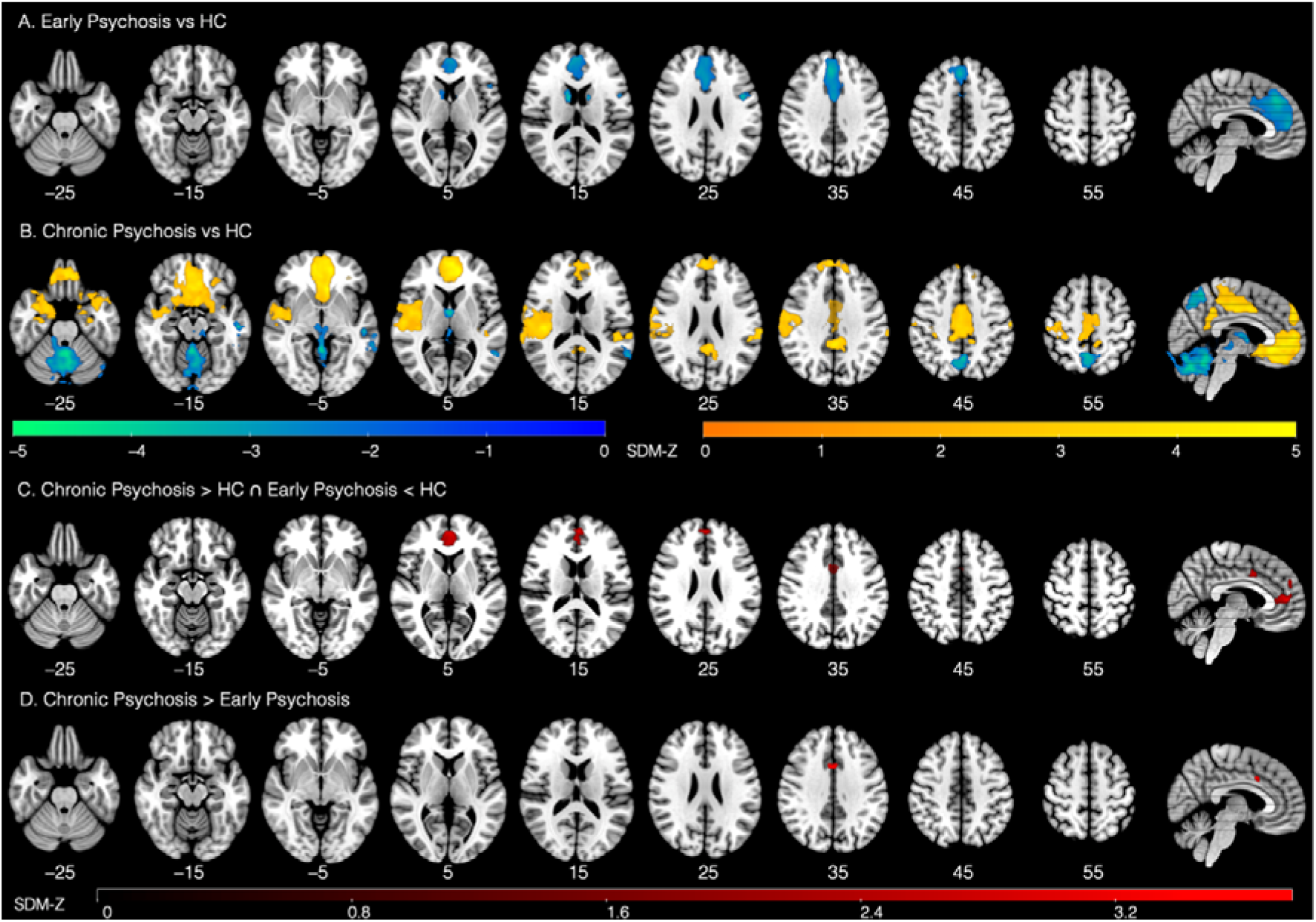
Meta-analytic findings in the early and chronic psychosis groups. The first 2 rows represent the significant clusters in the early (A) and chronic (B) psychosis meta-analyses. Only clusters larger than 10 voxels with *pFWE* < .05 are shown. C – the significant clusters identified in the disjunction analysis. D – significant cluster identified in the meta-comparison. The map was thresholded at p(uncorrected) < .0005. All sagittal section images are shown at x = 5.

**Table 5.** Peak coordinates of activation in the meta-analyses in early and chronic psychosis.

| Contrast | Cluster | Voxels | Grey matter regions | Highest peak<br>(MNI: x, y, z) | SDM-Z | *pFWE* |
| --- | --- | --- | --- | --- | --- | --- |
| *Early Psychosis > HC* | *No significant clusters* |  |  |  |  |  |
| *Early Psychosis < HC* | 1 | 3087 | Bilateral MSFG, ACC | 2, 32, 44 | -3.828 | <.001 |
|  | 2 | 144 | R Caudate nucleus | 10, 6, 10 | -4.023 | 0.007 |
|  | 3 | 103 | L IFG, opercular | -44, 4, 26 | -3.564 | 0.02 |
|  | 4 | 48 | L Caudate nucleus | -10, 6, 10 | -3.817 | 0.02 |
|  | 5 | 36 | L IFG, triangular | -50, 18, 8 | -3.44 | 0.036 |
| *Chronic Psychosis > HC* | 1 | 12561 | Bilateral ACC, MSFG, R Insula, Hippocampus, Striatum, R STG | -10, 44, 6 | 5.465 | <.001 |
|  | 2 | 3685 | Bilateral Median, Posterior cingulate | 0, -42, 38 | 4.701 | 0.004 |
|  | 3 | 478 | L STG | -38, -30, 12 | 3.738 | 0.018 |
|  | 4 | 51 | L Supramarginal gyrus | -58, -20, 42 | 3.939 | 0.044 |
| *Chronic Psychosis < HC* | 1 | 4652 | Cerebellum, thalamus | 0, -54, -26 | -4.661 | <.001 |
|  | 2 | 735 | Bilateral Precuneus | -2, -60, 44 | -3.849 | 0.01 |
|  | 3 | 557 | L Middle temporal gyrus | -54, -28, -10 | -4.309 | 0.01 |
|  | 4 | 179 | Thalamus | 2, -8, 8 | -4.072 | 0.014 |

*Abbreviations:* ACC – Anterior Cingulate Cortex, *FWE* – family-wise error, *HC* – healthy controls, *IFG* – inferior frontal gyrus, *L* – left, *MNI* – Montreal Neuroimaging Institute, *MSFG* – medial superior frontal gyrus, *R* – right, *STG* – superior temporal gyrus.

At the chronic psychosis stage, four significant clusters of hypoactivation relative to controls were identified in the cerebellum and thalamus, bilateral precuneus and left middle temporal gyrus, alongside four clusters of hyperactivation in the medial frontal cortex, cingulum, insula, ventral striatum, hippocampus, superior temporal and supramarginal gyri (Table 5; Figure 2B).Detailed cluster information for the early and chronic psychosis meta-analyses is presented in Tables S3-S4.

### 3.3. Conjunction analyses and meta-comparisons

Conjunction analyses were run to reveal overlaps in activity between the early and chronic psychosis group (vs respective HC groups), since the meta-analysis of the at-risk group did not yield significant differences at the chosen corrected threshold. No significantly overlapping regions of similar activation between the early psychosis and chronic psychosis groups were revealed.

Two regions of distinct activation patterns were revealed: the ACC (x=–2, y=50, z=8; 566 voxels; *pFWE*<.001) and middle cingulate (x=0, y=4, z=38; 107 voxels; *pFWE*<.002) (Figure 2C). The latter was also revealed in the meta-comparison, having significant hyperactivation in chronic psychosis, compared to early psychosis (x=4, y=12, z=34; *p_uncorr_*<.0005; SDM-*Z*=3.8; Figure 2D). Furthermore, we observed significant hypoactivation of the right caudate in early psychosis compared to the at-risk group (x=10, y=8, z=10; *p_uncorr_*<.0005; SDM-*Z*=3.48; Figure S2).

### 3.4. Meta-regressions

Meta-regressions revealed significant linear effects sex distribution in case-control proportion on WM-related brain activation in 3 clusters (Figure S3). A higher proportion of male participants in the psychosis group (compared to the control group) was associated with hypoactivation in the bilateral cerebellum (x=±28, y=–68/–62, z=–36, *p_uncorr_* < .0005), and hyperactivation in the superior medial frontal gyrus (x=8, y=56, z=18, SDM-*Z=*3.75, *p_uncorr_* < .001). No significant linear effects of sex distribution differences were found in the early psychosis or at-risk groups.

Finally, meta-regressions comprising all studies across the early and chronic psychosis stages revealed significant linear effects of antipsychotic dose and duration of illness on activation in the ACC. Both predictors revealed a positive association with ACC activation, with a higher degree of ACC activation in psychosis being associated with higher antipsychotic dose (Figure 3A), and longer duration of illness (Figure 3B).

**Figure 3.**
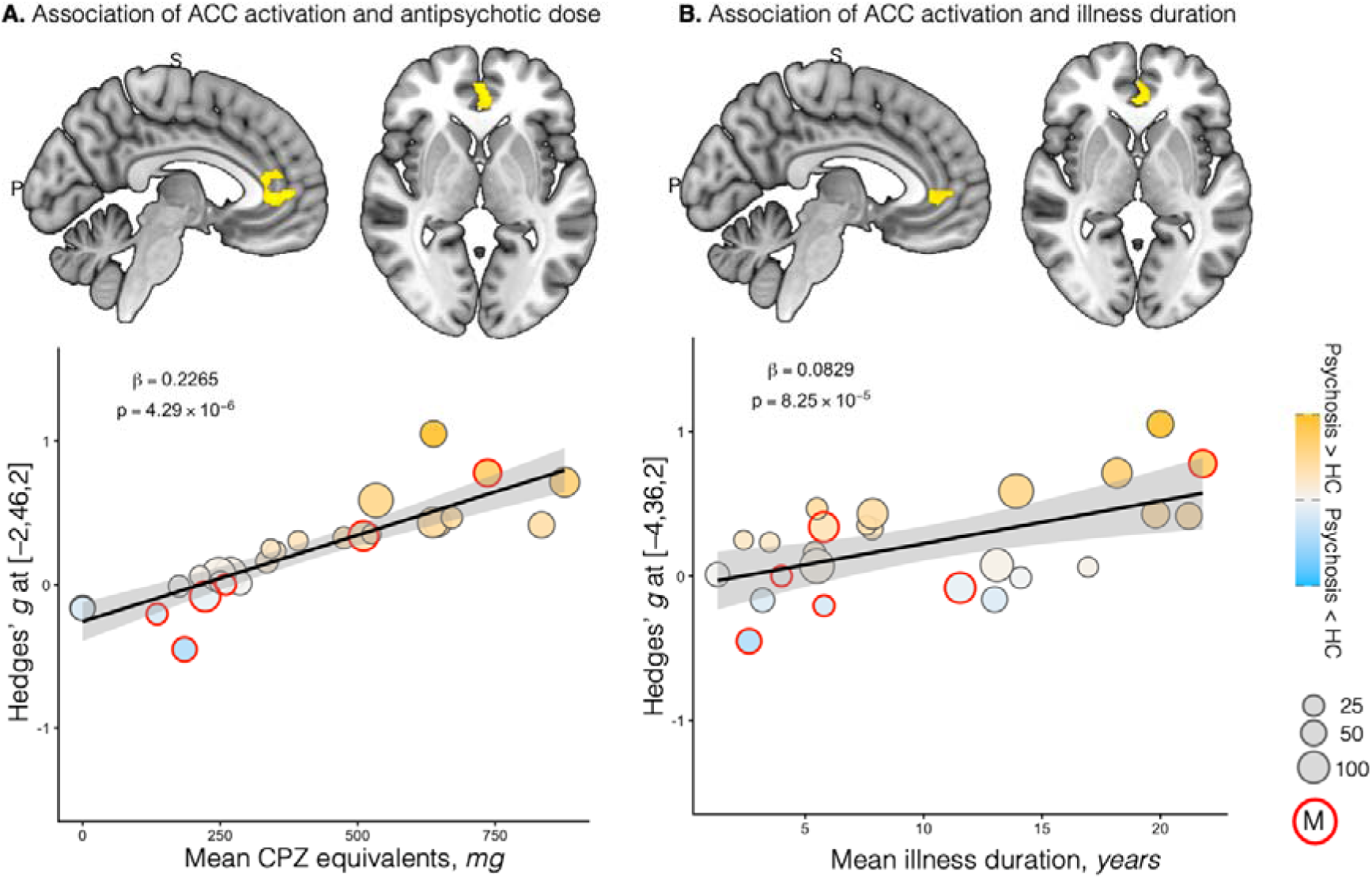
Meta-regression results showing significant linear effects of antipsychotic dose (A) and illness duration (B) on ACC activation compared to controls. ACC – anterior cingulate cortex, HC – healthy controls. PS – psychosis, CPZ – chlorpromazine, M – statistical maps; bubbles that are encompassed by a red circle represent effect sizes from studies where statistical maps were available. Clusters are shown at p <.0005 uncorrected for display purposes.

### 3.5. Sensitivity analyses

Jack-knife sensitivity analyses were conducted for the early and chronic psychosis groups. In the early psychosis group, the most robust clusters were in the ACC and right caudate, significant in 3/5 iterations (Table S5). For the chronic psychosis group, all clusters were replicated in at least 25/30 iterations (Table S6), the most robust clusters were in the cerebellum (30/30 iterations), ACC/superior medial frontal gyrus (29/30 iterations) and middle/posterior cingulate cortex (29/30 iterations). The findings from the sensitivity analyses where no covariates were included are shown in Tables S7-S8. The heterogeneity index (*I^2^)* was low across all analyses and no significant publication bias was detected in meta-bias tests (Tables S5-S6).

## 4. Discussion

This systematic review and meta-analysis revealed distinct patterns of WM-related brain activation across the early and chronic stages of psychosis, suggesting that WM-related fMRI alterations represent a state-dependent, rather than a trait, biomarker. These patterns were found in regions typically associated with the default mode and salience networks, with the most extensive alterations observed in the chronic psychosis stage. The ACC emerged as a hypoactive region in early psychosis, overactive in chronic psychosis, and its activation was associated with illness duration and antipsychotic dose. No statistically significant clusters emerged in the at-risk stage at a corrected threshold.

### 4.1. Chronic psychosis stage

The meta-analysis revealed hyperactivation in chronic psychosis compared to HC in regions that are core nodes of the default mode network (Foster and Koslov, 2025; Whitfield-Gabrieli and Ford, 2012), specifically the posterior cingulate cortex, medial prefrontal cortex, and the hippocampus. As these regions are typically deactivated during WM (Mencarelli et al., 2019; Wang et al., 2019), these findings may rather reflect hypo-deactivations. Abnormal suppression of the default mode network is associated with an unsuccessful switch from the internal attentional focus to externally-guided attention (Leech et al., 2011), and is a well-known fMRI signature in psychosis (Whitfield-Gabrieli and Ford, 2012).

In addition, we found WM-related brain hyperactivations or hypo-deactivations in the ACC and the insula, key hubs of the salience network, previously shown to be atypical in psychosis (Palaniyappan and Liddle, 2012). These findings suggest suboptimal allocation of resources to salient stimuli in cognitively demanding tasks, and altered regulation of large-scale functional networks (Chen et al., 2016; Goulden et al., 2014; Palaniyappan and Liddle, 2012).

Further significant effects comprised hypoactivations in n-back task-positive regions (Mencarelli et al., 2019; Wang et al., 2019), such as the precuneus, middle temporal gyrus, cerebellum and thalamus, which have been linked to cognitive dysfunction in schizophrenia (Cui et al., 2018; Dadario and Sughrue, 2023; Faris et al., 2024; Pergola et al., 2015). Notably, a distinct sub-region of the precuneus contributed to the posterior cingulate hypo-deactivation cluster, indicating functional heterogeneity across precuneus sub-regions, as suggested previously (Dadario and Sughrue, 2023). Supporting this, Utevsky et al (2014) found a dual role of the precuneus, showing opposing activity patterns when it is involved in the DMN (rest > task) versus the frontoparietal (task > rest) network.

Our meta-analytic findings in chronic psychosis replicate prior meta-analytic evidence of alterations in the ACC, posterior cingulate, and orbitofrontal cortices (Glahn et al., 2005; Wang et al., 2021; Wu and Jiang, 2020), thalamus (Andrews et al., 2006), and cerebellum (Bernard and Mittal, 2015; Yao et al., 2024), but contrast previous findings of insular hypoactivation (Wu and Jiang, 2020).

### 4.2. Early psychosis stage

In contrast with chronic psychosis, we did not find any clusters of WM-related hyperactivations (or hypo-deactivations) in early psychosis. We did observe widespread hypoactivations in the dorsal and pregenual ACC, consistent with its role in cognitive control and monitoring of salient information (Heilbronner and Hayden, 2016). Opposite effects were observed in the pregenual ACC across early and chronic psychosis stages, where we also found a linear effect of illness duration and antipsychotic dose. In the same ACC subregion, Radua et al (2012a) found reduced grey matter volumes in medicated patients with FEP, compared to non-medicated patients. These results suggest a potential of WM-related activation in the ACC as a neuroimaging biomarker for psychosis staging.

Furthermore, we found significant hypoactivation in the bilateral caudate nuclei in early psychosis, which were neither hyper-nor hypoactive in chronic psychosis Previous longitudinal studies found a normalisation of striatal hypoactivation over time in individuals with FEP, typically associated with antipsychotic treatment (Reske et al., 2007; Wulff et al., 2020). However, our study did not find a significant linear effect of antipsychotic dose, nor illness duration on striatal activity during WM.

### 4.3. At-risk for psychosis stage

The at-risk stage meta-analysis yielded no significant clusters after correction. This finding contrasts previous meta-analyses, which found significant WM-related frontoparietal alterations in in FHR (Zhang et al., 2016) and CHR-P (Dutt et al., 2015; Yao et al., 2024) separately. This lack of consensus is likely explained by the inclusion of ROI or masked findings, studies using other WM tasks, and less stringent statistical thresholds in previous meta-analyses. At a similar threshold as used previously (Yao et al., 2024), we did observe hyperactivation in the right middle frontal gyrus, consistent with prior meta-analytic evidence in FHR (Zhang et al., 2016). The lack of robust effects in this at-risk stage can also be explained by smaller effect sizes (Chan et al., 2011) and high heterogeneity, consistent with emerging evidence of distinct biotypes in CHR-P depending on fMRI activity (Tang et al., 2025). Alternatively, functional brain alterations may be distributed across different regions within the same large-scale networks (Darby et al., 2019; Makhlouf et al., 2025), and detecting these networks would require evidence from more studies. Future research could explicitly investigate the involvement of established networks in WM-related fMRI activation in both clinical and subclinical groups.

### 4.4 Strengths and limitations

This meta-analysis provides an updated synthesis of the WM-related fMRI literature across at-risk and clinical stages of psychosis. To our knowledge, this is the first meta-analysis to combine statistical maps with peak coordinates to examine stage-specific WM-related brain activity. To reduce heterogeneity and increase specificity, this meta-analysis focused on a single, widely used WM paradigm, applied stringent statistical thresholds, explored the effects of antipsychotic dose and illness duration, and excluded task contrasts reflecting non-WM processes. We identified a common practice in the fMRI literature involving the use of task activation masks derived from the same sample when performing second-level analyses. This approach, similar to ROI analyses (Müller et al., 2017), may bias meta-analytic results by not accounting for the whole brain, increasing the risk of false positives when cluster-extent thresholding is used (Eklund et al., 2016).

Our study has several limitations. We did not account for potential effects of substance use or acute symptom severity, which may influence fMRI activity (Weinberger and Radulescu, 2016). In addition, study-level differences in methodology, such as scanner field strength, smoothing kernel size, preprocessing and analysis software, and covariate selection, were not modelled in the analysis, limiting interpretability to an extent.

### 4.5. Conclusions

The present meta-analysis indicates that WM-related brain activity differs across stages of psychosis. Furthermore, we found stage-dependent alterations in the ACC (hypoactivation at early stages, hyperactivation at chronic stages) which were associated with antipsychotic dose and illness duration and are relevant to understanding cognitive symptoms and neural correlates of disease progression. Future studies may focus on investigating the differences between WM-specific and disease-specific alterations in psychosis, including examining the effects of antipsychotic type or disease status (relapse/remission) prospectively, to elucidate whether WM dysfunction may signal progression from early stages to overt illness, and inform stage-tailored intervention strategies.

## Declaration of generative AI and AI-assisted technologies in the writing process

During the preparation of this work the authors used ChatGPT 5 in order to ensure clarity and flow. After using this tool, the authors reviewed and edited the content as needed and take full responsibility for the content of the published article.

## Data Availability

All data produced in the present study are available upon reasonable request to the authors.

## Acknowledgement

We would like to thank all the researchers who kindly contributed statistical maps for the meta-analysis. We would also like to acknowledge Dr Abigail Gee and Nikita Catalina Julius, who provided informal feedback and helpful insights on diagnostic categories to help improve this work.

