## Supplement for "Neurofunctional correlates of Working Memory across Psychosis Stages: A Systematic Review and Meta-analysis"

**Neurofunctional correlates of working memory across the psychosis spectrum: A Systematic Review with Meta-analysis**

**Supplement**

### **Supplementary Methods**

#### ***Search terms used on Pubmed***

“((psychosis[Title/Abstract]) OR (schizophren*[Title/Abstract]) OR (SSD[Title/Abstract]) OR (schizoaffective[Title/Abstract]) OR (Clinical high-risk[Title/Abstract]) OR (ultra high-risk[Title/Abstract]) OR (familial high-risk[Title/Abstract]) OR (genetic high-risk[Title/Abstract]) OR (((siblings[Title/Abstract]) OR (offspring*[Title/Abstract]) OR (relative*[Title/Abstract])) AND (schizophren*[Title/Abstract]) ) OR (at-risk mental state[Title/Abstract]) OR (first-episode schizophrenia[Title/Abstract])OR (first-episode psychosis[Title/Abstract]) OR (FEP[Title/Abstract]) OR (Brief intermittent psychotic episode[Title/Abstract]) OR (schizotyp*[Title/Abstract]) OR (prodromal psychosis[Title/Abstract]) OR ((psychosis[Title/Abstract] OR psychotic[Title/Abstract] OR schizophren*[Title/Abstract]) AND (relatives[Title/Abstract] OR siblings[Title/Abstract] OR prodrome[Title/Abstract]))) AND ((functional magnetic resonance imaging[Title/Abstract]) OR (fmri[Title/Abstract]) OR (task-based neuroimaging[Title/Abstract]) OR (BOLD-fMRI[Title/Abstract]) OR (functional neuroimaging[Title/Abstract])) AND ((working memory task[Title/Abstract]) OR (working memory paradigm[Title/Abstract]) OR (-back[Title/Abstract]) OR (sternberg item recognition[Title/Abstract]) OR (SIRP[Title/Abstract]) OR (sternberg task[Title/Abstract]) OR (CPT[Title/Abstract])) NOT review”

#### ***Abstract and full text screening***

The screening of abstracts and full texts was performed by 2 reviewers (MMA and LBB) using the Rayyan platform (rayyan.ai). Both reviewers were blinded to each other’s decisions and followed the same list of eligibility criteria. At the abstract screening stage, the studies considered for inclusion were (1) observational studies using task-based fMRI that explored the difference in brain activation in psychosis stages(schizophrenia, schizo-affective disorder, first-episode psychosis) or at-risk groups(clinical high-risk for psychosis, familial high risk for psychosis) vs healthy controls while performing a the n-back task, (2) intervention studies that reported baseline measures of these differences, and (3) genetic analysis studies that reported differences between a psychosis stage/at-risk group (or multiple) and a healthy control group.

Once all abstracts had been screened, the reviewers were unblinded and joined a meeting to bring forward arguments for inclusion/exclusion when decisions were discrepant, until a consensus was reached on the final list of abstracts. When a decision could not be reached, a 3^rd^ investigator (MM or GM) was consulted. For full text screening, all publication PDFs were uploaded onto Rayyan, and the blinded screening process was repeated. Supplementary materials were screened where the studies did not report sufficient information for screening in the main text.

#### ***Data extraction***

Data extraction was performed by combining manual extraction by one of the reviewers (MMA) and AI-assisted extraction using a custom Generative Pre-Trained Transformer (GPT) *(openAI.com)* designed for this purpose. The extracted data included the following: ***paper information*** (authors, year of publication), ***demographics*** (country where the participants were recruited, populations studied, number of participants, mean age and sex distribution in each group, handedness, medication status, including antipsychotic and antidepressant use, dose of antipsychotic in chlorpromazine (CPZ) equivalents, illness duration, age of onset, cannabis use), ***methodology*** (type of scanner and field strength, WM paradigm, preprocessing and analysis software, main contrast for analysis and other contrasts if reported, normalisation template, smoothing gaussian kernel size (in full-width half maximum), the use of whole-brain vs. ROI findings, multiple comparison corrections, corrected and uncorrected thresholds applied, whether participants were matched for task performance, or trained in the task prior to testing), and ***results*** (peak coordinates of activation differences between population of interest and controls, activations, deactivations or non-significant findings per region in studies using ROI approaches or that did not use the n-back task). When a study reported coordinates derived from multiple contrasts (e.g. 2-back > 0-back, 2-back > 1-back), the peak coordinates were extracted from either the higher memory load or from the contrast that revealed significant corrected findings. After data extraction, the peak coordinates from each study were checked at least twice.

#### ***Meta-regressions***

Meta-regressions explored the effects of sex distribution, antipsychotic dose, and illness duration. The sex distribution variable was calculated by dividing the proportion of males in the cases group by the proportion of males in the control group, so that a value of 1 would indicate that the groups were matched perfectly for sex. For the meta-regression exploring effects of antipsychotic dose, mean CPZ equivalents(per dataset) was used as a predictor variable. For the meta-regression exploring the effects of illness duration, the mean duration of illness (years) was used as a predictor variable. In studies where illness duration was reported in months, we divided the reported mean number of months by 12 to convert it to years. Where age of onset was reported, but not illness duration, we used an approximate value by subtracting mean age at onset from mean age (current) to account for illness duration.

### **Supplementary tables and figures**

###### Table S1 – PRISMA checklist(Page et al., 2021)

| **Section and Topic** | **Item #** | **Checklist item** | **Location where item is reported** |
| --- | --- | --- | --- |
| **TITLE** | | |  |
| Title | 1 | Identify the report as a systematic review. | Page 1 |
| **ABSTRACT** | | |  |
| Abstract | 2 | See the PRISMA 2020 for Abstracts checklist. | Page 3-4 |
| **INTRODUCTION** | | |  |
| Rationale | 3 | Describe the rationale for the review in the context of existing knowledge. | Page 5-6 |
| Objectives | 4 | Provide an explicit statement of the objective(s) or question(s) the review addresses. | Page 6 |
| **METHODS** | | |  |
| Eligibility criteria | 5 | Specify the inclusion and exclusion criteria for the review and how studies were grouped for the syntheses. | Pages 6-7 |
| Information sources | 6 | Specify all databases, registers, websites, organisations, reference lists and other sources searched or consulted to identify studies. Specify the date when each source was last searched or consulted. | Page 6, Supplement page 2 |
| Search strategy | 7 | Present the full search strategies for all databases, registers and websites, including any filters and limits used. | Supplement page 2 |
| Selection process | 8 | Specify the methods used to decide whether a study met the inclusion criteria of the review, including how many reviewers screened each record and each report retrieved, whether they worked independently, and if applicable, details of automation tools used in the process. | Page 6, Supplement page 2-3 |
| Data collection process | 9 | Specify the methods used to collect data from reports, including how many reviewers collected data from each report, whether they worked independently, any processes for obtaining or confirming data from study investigators, and if applicable, details of automation tools used in the process. | Supplement page 2-4 |
| Data items | 10a | List and define all outcomes for which data were sought. Specify whether all results that were compatible with each outcome domain in each study were sought (e.g. for all measures, time points, analyses), and if not, the methods used to decide which results to collect. | Supplement page 3 |
|  | 10b | List and define all other variables for which data were sought (e.g. participant and intervention characteristics, funding sources). Describe any assumptions made about any missing or unclear information. | Supplement page 3 |
| Study risk of bias assessment | 11 | Specify the methods used to assess risk of bias in the included studies, including details of the tool(s) used, how many reviewers assessed each study and whether they worked independently, and if applicable, details of automation tools used in the process. | Page 8 |
| Effect measures | 12 | Specify for each outcome the effect measure(s) (e.g. risk ratio, mean difference) used in the synthesis or presentation of results. | Page 7 |
| Synthesis methods | 13a | Describe the processes used to decide which studies were eligible for each synthesis (e.g. tabulating the study intervention characteristics and comparing against the planned groups for each synthesis (item #5)). | Supplement page 2-3 |
|  | 13b | Describe any methods required to prepare the data for presentation or synthesis, such as handling of missing summary statistics, or data conversions. | Page 7-8, Supplement page 3-4 |
|  | 13c | Describe any methods used to tabulate or visually display results of individual studies and syntheses. | Page 6-8, Figure and Table legends. |
|  | 13d | Describe any methods used to synthesize results and provide a rationale for the choice(s). If meta-analysis was performed, describe the model(s), method(s) to identify the presence and extent of statistical heterogeneity, and software package(s) used. | Pages 6 and 7 |
|  | 13e | Describe any methods used to explore possible causes of heterogeneity among study results (e.g. subgroup analysis, meta-regression). | Page 8 |
|  | 13f | Describe any sensitivity analyses conducted to assess robustness of the synthesized results. | Page 8 |
| Reporting bias assessment | 14 | Describe any methods used to assess risk of bias due to missing results in a synthesis (arising from reporting biases). | Page 8 |
| Certainty assessment | 15 | Describe any methods used to assess certainty (or confidence) in the body of evidence for an outcome. | Not applicable |
| **RESULTS** | | |  |
| Study selection | 16a | Describe the results of the search and selection process, from the number of records identified in the search to the number of studies included in the review, ideally using a flow diagram. | Page 8 |
|  | 16b | Cite studies that might appear to meet the inclusion criteria, but which were excluded, and explain why they were excluded. | Page 8 |
| Study characteristics | 17 | Cite each included study and present its characteristics. | eTables 3-6 |
| Risk of bias in studies | 18 | Present assessments of risk of bias for each included study. | eTables 3-6 |
| Results of individual studies | 19 | For all outcomes, present, for each study: (a) summary statistics for each group (where appropriate) and (b) an effect estimate and its precision (e.g. confidence/credible interval), ideally using structured tables or plots. | Not applicable |
| Results of syntheses | 20a | For each synthesis, briefly summarise the characteristics and risk of bias among contributing studies. | Table 1 |
|  | 20b | Present results of all statistical syntheses conducted. If meta-analysis was done, present for each the summary estimate and its precision (e.g. confidence/credible interval) and measures of statistical heterogeneity. If comparing groups, describe the direction of the effect. | Page 9 |
|  | 20c | Present results of all investigations of possible causes of heterogeneity among study results. | Page 9-10 |
|  | 20d | Present results of all sensitivity analyses conducted to assess the robustness of the synthesized results. | Page 9-10 |
| Reporting biases | 21 | Present assessments of risk of bias due to missing results (arising from reporting biases) for each synthesis assessed. | Not applicable |
| Certainty of evidence | 22 | Present assessments of certainty (or confidence) in the body of evidence for each outcome assessed. | Not applicable |
| **DISCUSSION** | | |  |
| Discussion | 23a | Provide a general interpretation of the results in the context of other evidence. | Page 10 |
|  | 23b | Discuss any limitations of the evidence included in the review. | Page 12 |
|  | 23c | Discuss any limitations of the review processes used. | Page 12 |
|  | 23d | Discuss implications of the results for practice, policy, and future research. | Pages 11 and 13 |
| **OTHER INFORMATION** | | |  |
| Registration and protocol | 24a | Provide registration information for the review, including register name and registration number, or state that the review was not registered. | Page 6 |
|  | 24b | Indicate where the review protocol can be accessed, or state that a protocol was not prepared. | Page 6 |
|  | 24c | Describe and explain any amendments to information provided at registration or in the protocol. | Described in protocol |
| Support | 25 | Describe sources of financial or non-financial support for the review, and the role of the funders or sponsors in the review. | Page 13 |
| Competing interests | 26 | Declare any competing interests of review authors. | Page 13 |
| Availability of data, code and other materials | 27 | Report which of the following are publicly available and where they can be found: template data collection forms; data extracted from included studies; data used for all analyses; analytic code; any other materials used in the review. | Page 13 |

Table S2 – Results of the meta-analyses in the FHR and CHR-P groups separately. Results are shown at uncorrected p-value < .005, cluster size > 10 voxels.

| **Contrast** | **Cluster number** | **Highest MNI peak** | **SDM-Z** | **p*_uncorr._*** | **Voxels** | **GM regions** |
| --- | --- | --- | --- | --- | --- | --- |
| FHR < HC | 1 | 12,-44,4 | -4.016 | 0.000029564 | 234 | Right lingual gyrus, BA 27 |
| CHR-P < HC | 1 | 4,18,38 | -3.044 | 0.001167715 | 43 | Right median cingulate / paracingulate gyri, BA 24 |
|  | 2 | -36,44,26 | -3.569 | 0.000178933 | 28 | Left middle frontal gyrus, BA 46 |
|  | 3 | 4,14,52 | -2.876 | 0.002014637 | 10 | Right supplementary motor area |
| CHR-P > HC | 1 | -8,6,-2 | 3.235 | 0.00060904 | 47 | Left striatum |

*Abbreviations: CHR-P –* clinical high-risk for psychosis, *FHR* – familial high-risk for psychosis, *HC –* healthy controls, *MNI* – Montreal Neuroimaging Institute, *GM* – grey matter, *BA* – Brodmann Area, *SDM* – seed-based d-mapping.

Table S3 - Highest peaks of activity in the early psychosis meta-analysis. The results are reported at *pFWE* <.05.

| **Contrast** | **Cluster** | **MNI coordinate** | **SDM-Z** | **pFWE** | **Region** |
| --- | --- | --- | --- | --- | --- |
| Early psychosis  < HC | 1 | 2,32,44 | -3.828 | 0.00099999 | Left superior frontal gyrus, medial |
|  |  | 4,34,38 | -3.797 | ~0 | Right superior frontal gyrus, medial, BA 32 |
|  |  | -2,50,8 | -3.4 | 0.00099999 | Left anterior cingulate / paracingulate gyri, BA 32 |
|  |  | 6,48,10 | -3.384 | 0.00099999 | Right anterior cingulate / paracingulate gyri, BA 32 |
|  |  | 0,46,8 | -3.378 | 0.00099999 | Left anterior cingulate / paracingulate gyri |
|  |  | 6,44,38 | -3.362 | 0.00099999 | Right superior frontal gyrus, medial, BA 9 |
|  |  | -2,50,12 | -3.329 | 0.00099999 | Left anterior cingulate / paracingulate gyri, BA 32 |
|  |  | 4,52,16 | -3.256 | 0.00099999 | Right superior frontal gyrus, medial, BA 32 |
|  |  | 0,4,38 | -3.15 | 0.00099999 | Left median cingulate / paracingulate gyri, BA 24 |
|  |  | 0,30,24 | -3.142 | 0.00099999 | Left anterior cingulate / paracingulate gyri |
|  |  | 0,26,26 | -3.121 | 0.00099999 | Left anterior cingulate / paracingulate gyri |
|  |  | -6,44,6 | -3.091 | 0.00099999 | Left anterior cingulate / paracingulate gyri, BA 32 |
|  |  | 10,34,22 | -3.08 | 0.00099999 | Right anterior cingulate / paracingulate gyri |
|  |  | 12,36,48 | -3.078 | 0.00099999 | Right superior frontal gyrus, medial, BA 9 |
|  |  | 8,34,48 | -3.063 | 0.00099999 | Corpus callosum |
|  |  | 0,16,36 | -3.017 | 0.00099999 | Left median cingulate / paracingulate gyri, BA 24 |
|  |  | 0,36,4 | -2.979 | 0.00099999 | Left anterior cingulate / paracingulate gyri, BA 25 |
|  |  | 0,42,26 | -2.964 | 0.00099999 | Left superior frontal gyrus, medial, BA 32 |
|  |  | 0,46,24 | -2.955 | 0.00099999 | Left superior frontal gyrus, medial, BA 32 |
|  |  | 2,34,8 | -2.824 | 0.00099999 | (undefined) |
|  |  | 0,38,18 | -2.734 | 0.00199997 | Left anterior cingulate / paracingulate gyri, BA 24 |
|  |  | -10,20,36 | -2.187 | 0.04699999 | Left median cingulate / paracingulate gyri, BA 32 |
|  | 2 | 10,6,10 | -4.023 | 0.00700003 | Right anterior thalamic projections |
|  |  | 10,6,14 | -4.015 | 0.00700003 | Right caudate nucleus |
|  | 3 | -44,4,26 | -3.564 | 0.01999998 | Left inferior frontal gyrus, opercular part, BA 44 |
|  |  | -48,6,22 | -3.444 | 0.02200002 | Left precentral gyrus, BA 44 |
|  | 4 | -10,6,10 | -3.817 | 0.02200002 | Left anterior thalamic projections |
|  |  | -12,4,16 | -3.669 | 0.023 | Left anterior thalamic projections |
|  | 5 | -50,18,8 | -3.44 | 0.03600001 | Left inferior frontal gyrus, triangular part, BA 48 |

*Abbreviations: BA* – Brodmann Area, *HC* – healthy controls*, pFWE* –family-wise error corrected p-value, *SDM* – seed-based d-mapping.

Table S4 – Highest peaks of activity for the chronic psychosis meta-analysis. The results are reported at *pFWE* <.05.

| **Contrast** | **Cluster** | **MNI coordinate** | **SDM-Z** | **pFWE** | **Region** |
| --- | --- | --- | --- | --- | --- |
| Chronic psychosis > HC | 1 | -10,44,6 | 5.465 | ~0 | Left anterior cingulate / paracingulate gyri, BA 32 |
|  |  | -8,26,-8 | 5.306 | 0.00099999 | Corpus callosum |
|  |  | 0,54,2 | 5.226 | 0.00099999 | Left superior frontal gyrus, medial |
|  |  | -10,46,-8 | 5.2 | 0.00099999 | Left superior frontal gyrus, medial orbital, BA 10 |
|  |  | -6,24,-4 | 5.192 | 0.00099999 | Corpus callosum |
|  |  | -2,44,8 | 5.138 | 0.00099999 | Left anterior cingulate / paracingulate gyri, BA 32 |
|  |  | -4,52,4 | 5.118 | 0.00099999 | Left superior frontal gyrus, medial, BA 10 |
|  |  | 6,30,-8 | 5.112 | 0.00099999 | Right anterior cingulate / paracingulate gyri, BA 11 |
|  |  | -6,48,6 | 5.105 | 0.00099999 | Left anterior cingulate / paracingulate gyri, BA 10 |
|  |  | -6,44,6 | 5.088 | 0.00099999 | Left anterior cingulate / paracingulate gyri, BA 32 |
|  |  | 40,-22,14 | 5.08 | 0.00099999 | Right heschl gyrus, BA 48 |
|  |  | -10,42,-20 | 4.931 | 0.00099999 | Left gyrus rectus, BA 11 |
|  |  | -2,26,-8 | 4.845 | 0.00099999 | Left anterior cingulate / paracingulate gyri, BA 11 |
|  |  | -2,24,-12 | 4.837 | 0.00099999 | Left olfactory cortex, BA 11 |
|  |  | 8,46,-6 | 4.789 | 0.00099999 | Right superior frontal gyrus, medial orbital, BA 10 |
|  |  | 6,42,-2 | 4.776 | 0.00099999 | Right anterior cingulate / paracingulate gyri, BA 10 |
|  |  | 8,30,-18 | 4.752 | 0.00099999 | Corpus callosum |
|  |  | 0,44,2 | 4.707 | 0.00099999 | Left anterior cingulate / paracingulate gyri |
|  |  | -2,42,-24 | 4.704 | 0.00099999 | Left gyrus rectus, BA 11 |
|  |  | 0,46,-22 | 4.523 | 0.00099999 | Left gyrus rectus, BA 11 |
|  |  | 4,34,-20 | 4.394 | 0.00099999 | Right gyrus rectus, BA 11 |
|  |  | -4,56,-4 | 4.384 | 0.00099999 | Left superior frontal gyrus, medial orbital, BA 10 |
|  |  | 6,14,-12 | 4.368 | 0.00099999 | Right striatum |
|  |  | -8,40,-2 | 4.366 | 0.00099999 | Left median network, cingulum |
|  |  | 4,54,30 | 4.362 | 0.00099999 | Right superior frontal gyrus, medial, BA 10 |
|  |  | 2,40,-4 | 4.339 | 0.00099999 | Left anterior cingulate / paracingulate gyri, BA 11 |
|  |  | -4,48,-6 | 4.326 | 0.00099999 | Left superior frontal gyrus, medial orbital, BA 10 |
|  |  | -6,12,-14 | 4.318 | 0.00099999 | Left striatum |
|  |  | -6,36,-12 | 4.292 | 0.00099999 | Left superior frontal gyrus, medial orbital, BA 11 |
|  |  | 56,-16,6 | 4.254 | 0.00099999 | Right superior temporal gyrus, BA 48 |
|  |  | 44,-4,6 | 4.228 | 0.00199997 | Right insula, BA 48 |
|  |  | 12,40,-22 | 4.203 | 0.00099999 | Right gyrus rectus, BA 11 |
|  |  | 12,36,-22 | 4.086 | 0.00099999 | Right gyrus rectus, BA 11 |
|  |  | 0,42,-16 | 4.072 | 0.00099999 | Left gyrus rectus, BA 11 |
|  |  | 8,36,-22 | 4.02 | 0.00099999 | Right gyrus rectus, BA 11 |
|  |  | 34,10,10 | 3.967 | 0.00199997 | Right insula, BA 48 |
|  |  | 38,-4,8 | 3.935 | 0.00199997 | Right insula, BA 48 |
|  |  | 26,-2,-22 | 3.86 | 0.00099999 | Right amygdala, BA 34 |
|  |  | 16,28,-20 | 3.761 | 0.00099999 | Right frontal orbito-polar tract |
|  |  | 66,-32,12 | 3.753 | 0.00400001 | Right superior temporal gyrus, BA 22 |
|  |  | 22,0,-26 | 3.74 | 0.00199997 | Right parahippocampal gyrus, BA 28 |
|  |  | 50,-22,50 | 3.707 | 0.02700001 | Right postcentral gyrus, BA 3 |
|  |  | 2,46,-12 | 3.679 | 0.00099999 | Right superior frontal gyrus, medial orbital, BA 11 |
|  |  | 34,0,-24 | 3.675 | 0.00199997 | Right amygdala, BA 36 |
|  |  | 36,4,-26 | 3.648 | 0.00199997 | (undefined), BA 38 |
|  |  | -38,2,-22 | 3.635 | 0.00400001 | (undefined), BA 38 |
|  |  | 18,18,-18 | 3.629 | 0.00199997 | Right gyrus rectus, BA 11 |
|  |  | -4,52,28 | 3.627 | 0.00099999 | Left superior frontal gyrus, medial, BA 10 |
|  |  | 50,-4,-4 | 3.622 | 0.00199997 | Right superior temporal gyrus, BA 48 |
|  |  | 58,-16,14 | 3.615 | 0.00199997 | Right rolandic operculum, BA 48 |
|  |  | -8,52,30 | 3.604 | 0.00099999 | Corpus callosum |
|  |  | 6,54,40 | 3.596 | 0.00099999 | Right superior frontal gyrus, medial, BA 9 |
|  |  | -18,20,-20 | 3.552 | 0.00099999 | Left inferior frontal gyrus, orbital part, BA 11 |
|  |  | 40,10,8 | 3.525 | 0.00400001 | Right inferior frontal gyrus, opercular part, BA 48 |
|  |  | 64,-18,32 | 3.5 | 0.00599998 | Right postcentral gyrus, BA 43 |
|  |  | 58,-10,-2 | 3.469 | 0.00199997 | Corpus callosum |
|  |  | 54,-10,36 | 3.461 | 0.00599998 | Right postcentral gyrus, BA 3 |
|  |  | 36,2,-32 | 3.436 | 0.00199997 | Right inferior network, inferior longitudinal fasciculus |
|  |  | 22,8,-20 | 3.425 | 0.00199997 | Right temporal pole, superior temporal gyrus, BA 34 |
|  |  | 66,-6,20 | 3.417 | 0.00700003 | Right postcentral gyrus, BA 43 |
|  |  | 32,-30,46 | 3.393 | 0.028 | Right superior longitudinal fasciculus II |
|  |  | 46,-6,-12 | 3.319 | 0.00199997 | Right inferior network, inferior longitudinal fasciculus |
|  |  | 58,-8,36 | 3.314 | 0.00599998 | Right postcentral gyrus, BA 3 |
|  |  | 38,-6,-8 | 3.31 | 0.00199997 | (undefined), BA 48 |
|  |  | 30,-14,-24 | 3.308 | 0.01800001 | Right median network, cingulum |
|  |  | -22,0,-18 | 3.302 | 0.00400001 | Left amygdala, BA 34 |
|  |  | -22,6,-14 | 3.292 | 0.00400001 | Left olfactory cortex, BA 48 |
|  |  | 22,10,-16 | 3.29 | 0.00199997 | Right olfactory cortex, BA 48 |
|  |  | -28,-12,-24 | 3.28 | 0.00700003 | Left median network, cingulum |
|  |  | 44,-10,34 | 3.265 | 0.00599998 | Right postcentral gyrus, BA 4 |
|  |  | 60,-4,32 | 3.257 | 0.00599998 | Right postcentral gyrus, BA 4 |
|  |  | 60,-20,34 | 3.255 | 0.00700003 | Right postcentral gyrus, BA 43 |
|  |  | 52,-2,10 | 3.248 | 0.00199997 | Right rolandic operculum, BA 48 |
|  |  | -40,8,-24 | 3.24 | 0.005 | Left temporal pole, superior temporal gyrus, BA 38 |
|  |  | -40,12,-24 | 3.236 | 0.005 | Left temporal pole, superior temporal gyrus, BA 38 |
|  |  | 64,-30,20 | 3.209 | 0.005 | Right superior temporal gyrus, BA 48 |
|  |  | 54,2,10 | 3.195 | 0.00400001 | Right rolandic operculum, BA 48 |
|  |  | 22,34,-10 | 3.187 | 0.01200002 | Right frontal orbito-polar tract |
|  |  | -18,0,-20 | 3.184 | 0.00400001 | Left amygdala, BA 34 |
|  |  | 40,-6,-12 | 3.152 | 0.00199997 | (undefined), BA 48 |
|  |  | 58,-26,36 | 3.13 | 0.00700003 | Right supramarginal gyrus, BA 2 |
|  |  | 64,0,18 | 3.129 | 0.00700003 | Right postcentral gyrus, BA 43 |
|  |  | -18,-6,-22 | 3.115 | 0.00400001 | Left hippocampus, BA 28 |
|  |  | 12,56,36 | 3.111 | 0.00099999 | Right superior frontal gyrus, medial, BA 9 |
|  |  | 34,-34,62 | 3.075 | 0.03299999 | Right postcentral gyrus, BA 4 |
|  |  | 50,-26,18 | 3.074 | 0.00199997 | Right rolandic operculum, BA 48 |
|  |  | 30,0,-42 | 3.046 | 0.00400001 | Right fusiform gyrus, BA 20 |
|  |  | -22,-4,-22 | 3.031 | 0.00400001 | Left amygdala, BA 28 |
|  |  | 54,-22,34 | 3.029 | 0.00700003 | Right superior longitudinal fasciculus III |
|  |  | 2,18,-4 | 3.02 | 0.00199997 | Right olfactory cortex, BA 25 |
|  |  | 32,-18,64 | 3.01 | 0.03299999 | Right precentral gyrus, BA 6 |
|  |  | 22,4,-38 | 3.008 | 0.00400001 | Right temporal pole, middle temporal gyrus, BA 36 |
|  |  | 60,4,6 | 2.993 | 0.00400001 | Right rolandic operculum, BA 48 |
|  |  | 58,8,-2 | 2.993 | 0.01200002 | Right temporal pole, superior temporal gyrus, BA 38 |
|  |  | 16,54,30 | 2.992 | 0.00199997 | Right superior frontal gyrus, dorsolateral, BA 9 |
|  |  | 32,-32,54 | 2.988 | 0.03299999 | Right postcentral gyrus, BA 3 |
|  |  | 66,-36,16 | 2.975 | 0.005 | Right superior temporal gyrus, BA 42 |
|  |  | 34,-14,64 | 2.974 | 0.03299999 | Right precentral gyrus, BA 6 |
|  |  | -26,18,-22 | 2.969 | 0.00199997 | Left inferior frontal gyrus, orbital part, BA 38 |
|  |  | -16,8,-16 | 2.947 | 0.00400001 | Left olfactory cortex, BA 11 |
|  |  | -32,16,-26 | 2.946 | 0.00400001 | Left temporal pole, superior temporal gyrus, BA 38 |
|  |  | 62,-24,38 | 2.939 | 0.00700003 | Right supramarginal gyrus, BA 1 |
|  |  | 16,50,34 | 2.921 | 0.00199997 | Right superior frontal gyrus, dorsolateral, BA 9 |
|  |  | 48,-10,16 | 2.916 | 0.00400001 | Right rolandic operculum, BA 48 |
|  |  | -26,38,-10 | 2.888 | 0.01099998 | Left middle frontal gyrus, orbital part, BA 11 |
|  |  | 26,10,-30 | 2.885 | 0.00400001 | Right parahippocampal gyrus, BA 38 |
|  |  | 28,14,-24 | 2.884 | 0.00400001 | Right inferior frontal gyrus, orbital part, BA 38 |
|  |  | 38,-24,58 | 2.881 | 0.03200001 | Right precentral gyrus, BA 4 |
|  |  | 38,-18,58 | 2.869 | 0.03200001 | Right precentral gyrus, BA 4 |
|  |  | 36,-10,-26 | 2.868 | 0.02200002 | Right inferior network, inferior longitudinal fasciculus |
|  |  | 34,10,-28 | 2.858 | 0.00400001 | Right temporal pole, superior temporal gyrus, BA 38 |
|  |  | 44,-20,38 | 2.853 | 0.00800002 | Right superior longitudinal fasciculus III |
|  |  | 60,2,-4 | 2.851 | 0.01200002 | Right superior temporal gyrus, BA 38 |
|  |  | -32,12,-26 | 2.849 | 0.00599998 | Left temporal pole, superior temporal gyrus, BA 38 |
|  |  | 16,12,-16 | 2.842 | 0.00400001 | Right olfactory cortex, BA 11 |
|  |  | 62,-24,12 | 2.831 | 0.00400001 | Right superior temporal gyrus, BA 22 |
|  |  | -34,16,-22 | 2.815 | 0.00400001 | Left temporal pole, superior temporal gyrus |
|  |  | 34,-24,60 | 2.813 | 0.03500003 | Right precentral gyrus, BA 4 |
|  |  | -14,40,42 | 2.801 | 0.01099998 | Left superior frontal gyrus, dorsolateral, BA 32 |
|  |  | -34,38,-8 | 2.797 | 0.01200002 | Left inferior frontal gyrus, orbital part, BA 47 |
|  |  | 26,42,-10 | 2.793 | 0.01200002 | Right inferior network, inferior fronto-occipital fasciculus |
|  |  | -42,-2,-36 | 2.781 | 0.01800001 | Left inferior temporal gyrus, BA 20 |
|  |  | 64,-14,16 | 2.756 | 0.00599998 | Right postcentral gyrus, BA 48 |
|  |  | 14,-4,-26 | 2.747 | 0.00599998 | (undefined) |
|  |  | 32,-20,60 | 2.746 | 0.03600001 | Right precentral gyrus, BA 6 |
|  |  | -4,54,38 | 2.744 | 0.00599998 | Left superior frontal gyrus, medial, BA 9 |
|  |  | -28,36,-14 | 2.743 | 0.01200002 | Left middle frontal gyrus, orbital part, BA 11 |
|  |  | 66,-20,8 | 2.736 | 0.00400001 | Right superior temporal gyrus, BA 22 |
|  |  | -26,44,-10 | 2.732 | 0.01200002 | Left middle frontal gyrus, orbital part, BA 11 |
|  |  | 46,-14,18 | 2.691 | 0.00400001 | Right rolandic operculum, BA 48 |
|  |  | -24,30,-14 | 2.671 | 0.01200002 | Left inferior frontal gyrus, orbital part, BA 11 |
|  |  | 50,-30,24 | 2.642 | 0.014 | Right supramarginal gyrus, BA 48 |
|  |  | 38,-12,20 | 2.626 | 0.00599998 | Right insula, BA 48 |
|  |  | -52,8,-26 | 2.604 | 0.01300001 | Left middle temporal gyrus, BA 21 |
|  |  | 56,-32,16 | 2.588 | 0.00800002 | Right superior temporal gyrus, BA 42 |
|  |  | 26,20,-22 | 2.586 | 0.01099998 | Right inferior frontal gyrus, orbital part, BA 38 |
|  |  | -38,-8,-30 | 2.578 | 0.04299998 | Left inferior network, inferior longitudinal fasciculus |
|  |  | -38,0,-32 | 2.568 | 0.01800001 | Left inferior network, inferior longitudinal fasciculus |
|  |  | 62,-18,24 | 2.568 | 0.00800002 | Right supramarginal gyrus, BA 48 |
|  |  | 38,22,-26 | 2.566 | 0.04299998 | Right temporal pole, superior temporal gyrus, BA 38 |
|  |  | -10,56,22 | 2.53 | 0.01099998 | Corpus callosum |
|  |  | 64,-30,36 | 2.529 | 0.01099998 | Right supramarginal gyrus, BA 2 |
|  |  | -38,6,-32 | 2.524 | 0.01099998 | Left inferior network, inferior longitudinal fasciculus |
|  |  | 64,-24,18 | 2.512 | 0.01300001 | Right supramarginal gyrus, BA 42 |
|  |  | -36,20,-24 | 2.471 | 0.01800001 | Left temporal pole, superior temporal gyrus, BA 38 |
|  |  | -50,14,-26 | 2.468 | 0.01300001 | Left temporal pole, middle temporal gyrus, BA 38 |
|  |  | 20,42,-14 | 2.357 | 0.01800001 | Right frontal orbito-polar tract |
|  |  | 56,-28,28 | 2.357 | 0.014 | Right supramarginal gyrus, BA 48 |
|  |  | -6,18,-22 | 2.356 | 0.01800001 | Left striatum |
|  |  | 40,-22,44 | 2.337 | 0.04799998 | Right hand middle U tract |
|  |  | -38,4,-38 | 2.332 | 0.014 | Left inferior temporal gyrus, BA 36 |
|  |  | 36,-22,44 | 2.312 | 0.04799998 | Right superior longitudinal fasciculus II |
|  |  | 46,-12,42 | 2.273 | 0.02600002 | Right precentral gyrus, BA 4 |
|  |  | 16,50,42 | 2.271 | 0.014 | Right superior frontal gyrus, dorsolateral, BA 9 |
|  |  | -34,12,-34 | 2.253 | 0.037 | Left temporal pole, middle temporal gyrus, BA 20 |
|  |  | 2,18,-22 | 2.247 | 0.037 | Right gyrus rectus, BA 11 |
|  |  | 24,14,-8 | 2.109 | 0.037 | Right lenticular nucleus, putamen, BA 11 |
|  | 2 | 0,-42,38 | 4.701 | 0.00400001 | Left median cingulate / paracingulate gyri |
|  |  | 2,-10,40 | 4.582 | 0.00199997 | Right median cingulate / paracingulate gyri, BA 23 |
|  |  | -4,0,38 | 4.526 | 0.00199997 | Left median cingulate / paracingulate gyri, BA 24 |
|  |  | -2,-44,30 | 4.495 | 0.00400001 | Left posterior cingulate gyrus, BA 23 |
|  |  | 4,2,40 | 4.258 | 0.00199997 | Right median cingulate / paracingulate gyri, BA 24 |
|  |  | 2,-20,42 | 4.075 | 0.00199997 | Right median cingulate / paracingulate gyri, BA 23 |
|  |  | -2,-12,40 | 4.052 | 0.00199997 | Left median cingulate / paracingulate gyri, BA 23 |
|  |  | 8,-16,40 | 4.037 | 0.00199997 | Right median cingulate / paracingulate gyri, BA 23 |
|  |  | 10,-22,42 | 3.782 | 0.00400001 | Right median cingulate / paracingulate gyri |
|  |  | 0,-26,42 | 3.748 | 0.00400001 | Left median cingulate / paracingulate gyri, BA 23 |
|  |  | 12,-34,46 | 3.684 | 0.00400001 | Right median cingulate / paracingulate gyri |
|  |  | 8,-30,60 | 3.578 | 0.00599998 | Corpus callosum |
|  |  | -4,-22,40 | 3.554 | 0.00400001 | Left median cingulate / paracingulate gyri, BA 23 |
|  |  | 14,-32,40 | 3.531 | 0.005 | Right median cingulate / paracingulate gyri |
|  |  | -8,-36,40 | 3.516 | 0.00599998 | Left median cingulate / paracingulate gyri |
|  |  | 0,-34,48 | 3.415 | 0.00400001 | Left median cingulate / paracingulate gyri |
|  |  | 16,-30,68 | 3.324 | 0.00599998 | Right precentral gyrus, BA 4 |
|  |  | 8,-18,52 | 3.285 | 0.00400001 | Corpus callosum |
|  |  | -6,-48,20 | 3.276 | 0.01099998 | Left median network, cingulum |
|  |  | -8,-8,40 | 3.18 | 0.00599998 | Left median cingulate / paracingulate gyri, BA 23 |
|  |  | -8,-40,52 | 3.081 | 0.01300001 | Left median cingulate / paracingulate gyri |
|  |  | -6,-6,54 | 3.07 | 0.00999999 | Corpus callosum |
|  |  | -8,-18,58 | 3.057 | 0.014 | Corpus callosum |
|  |  | -8,-8,48 | 3.032 | 0.009 | Left median cingulate / paracingulate gyri |
|  |  | -2,-30,62 | 3.007 | 0.01200002 | Left paracentral lobule |
|  |  | 2,-48,18 | 2.986 | 0.014 | (undefined) |
|  |  | 8,-36,62 | 2.96 | 0.00700003 | Corpus callosum |
|  |  | -10,-52,24 | 2.953 | 0.01300001 | Left precuneus, BA 23 |
|  |  | -12,-50,28 | 2.901 | 0.01200002 | Left median network, cingulum |
|  |  | -6,-10,58 | 2.876 | 0.01099998 | Left supplementary motor area, BA 6 |
|  |  | 2,-26,62 | 2.833 | 0.01099998 | Right supplementary motor area, BA 4 |
|  |  | 8,-44,58 | 2.807 | 0.01800001 | Right precuneus |
|  |  | 2,-22,56 | 2.801 | 0.01099998 | Right supplementary motor area, BA 4 |
|  |  | -14,-28,40 | 2.788 | 0.01300001 | Left median cingulate / paracingulate gyri |
|  |  | -6,-40,60 | 2.692 | 0.014 | Left precuneus, BA 5 |
|  |  | 10,-36,74 | 2.625 | 0.02999997 | Right paracentral lobule, BA 4 |
|  |  | 14,-34,72 | 2.601 | 0.02999997 | Right postcentral gyrus, BA 4 |
|  |  | -6,-46,60 | 2.532 | 0.03799999 | Left precuneus, BA 5 |
|  |  | 8,-4,56 | 2.481 | 0.03200001 | Right supplementary motor area, BA 6 |
|  |  | 8,-28,74 | 2.433 | 0.03799999 | Right paracentral lobule, BA 4 |
|  |  | 10,-40,40 | 2.353 | 0.03299999 | Right median cingulate / paracingulate gyri |
|  |  | -8,-48,10 | 2.35 | 0.02700001 | Left precuneus, BA 29 |
|  |  | -4,-24,58 | 2.345 | 0.02100003 | Left paracentral lobule, BA 4 |
|  |  | 2,-8,60 | 2.196 | 0.03899998 | Right supplementary motor area, BA 6 |
|  | 3 | -38,-30,12 | 3.738 | 0.01800001 | Left arcuate network, posterior segment |
|  |  | -44,-32,10 | 3.709 | 0.01800001 | Left superior temporal gyrus, BA 41 |
|  |  | -50,-34,18 | 3.683 | 0.01800001 | Left superior temporal gyrus, BA 41 |
|  |  | -60,-32,16 | 3.619 | 0.02700001 | Left superior temporal gyrus, BA 42 |
|  |  | -64,-26,24 | 3.617 | 0.01899999 | Left supramarginal gyrus, BA 48 |
|  |  | -64,-28,32 | 3.578 | 0.01899999 | Left supramarginal gyrus, BA 2 |
|  |  | -62,-32,12 | 3.477 | 0.02700001 | Left superior temporal gyrus, BA 22 |
|  |  | -60,-24,20 | 3.464 | 0.02999997 | Left supramarginal gyrus, BA 48 |
|  |  | -64,-32,30 | 3.407 | 0.01899999 | Left supramarginal gyrus, BA 2 |
|  |  | -60,-34,26 | 3.309 | 0.02100003 | Left supramarginal gyrus, BA 48 |
|  |  | -52,-36,24 | 3.266 | 0.02100003 | Left supramarginal gyrus, BA 48 |
|  |  | -60,-32,22 | 3.178 | 0.02200002 | Left superior temporal gyrus, BA 48 |
|  |  | -64,-28,14 | 3.044 | 0.03899998 | Left superior temporal gyrus, BA 42 |
|  |  | -34,-22,6 | 2.67 | 0.04299998 | Left heschl gyrus, BA 48 |
|  | 4 | -58,-20,42 | 3.939 | 0.03100002 | Left supramarginal gyrus, BA 3 |
|  |  | -52,-20,38 | 3.427 | 0.04100001 | Left inferior parietal (excluding supramarginal and angular) gyri, BA 3 |
| Chronic psychosis < HC | 1 | 0,-54,-26 | -4.661 | ~0 | (undefined) |
|  |  | 4,-54,-26 | -4.626 | ~0 | (undefined) |
|  |  | -4,-62,-24 | -4.356 | 0.00099999 | Cerebellum, vermic lobule VI |
|  |  | 6,-68,-32 | -4.328 | 0.00099999 | Right cerebellum, crus II |
|  |  | 6,-72,-32 | -4.275 | 0.00099999 | Right cerebellum, crus II |
|  |  | 0,-48,-6 | -4.249 | 0.00099999 | Cerebellum, vermic lobule IV / V |
|  |  | 4,-50,-16 | -4.248 | 0.00099999 | Cerebellum, vermic lobule IV / V |
|  |  | -4,-68,-28 | -4.12 | 0.00099999 | Cerebellum, vermic lobule VII |
|  |  | -2,-68,-24 | -4.056 | 0.00099999 | Cerebellum, vermic lobule VI |
|  |  | 2,-56,-36 | -4.055 | 0.00099999 | Cerebellum, vermic lobule IX |
|  |  | -4,-70,-32 | -3.965 | 0.00099999 | Left cerebellum, crus II |
|  |  | -8,-66,-34 | -3.861 | 0.00099999 | (undefined) |
|  |  | 6,-74,-18 | -3.824 | 0.00099999 | Cerebellum, vermic lobule VI |
|  |  | 0,-56,-8 | -3.78 | 0.00099999 | Cerebellum, vermic lobule IV / V |
|  |  | 20,-64,-34 | -3.763 | 0.00099999 | (undefined) |
|  |  | -4,-64,-12 | -3.646 | 0.00099999 | Left cerebellum, hemispheric lobule VI |
|  |  | 14,-64,-34 | -3.644 | 0.00099999 | (undefined) |
|  |  | -4,-56,-10 | -3.61 | 0.00099999 | Left cerebellum, hemispheric lobule IV / V, BA 18 |
|  |  | 0,-76,-20 | -3.608 | 0.00099999 | Cerebellum, vermic lobule VII |
|  |  | 10,-68,-24 | -3.549 | 0.00099999 | Right cerebellum, hemispheric lobule VI |
|  |  | 12,-44,-28 | -3.431 | 0.00099999 | (undefined) |
|  |  | 18,-62,-24 | -3.421 | 0.00099999 | Right cerebellum, hemispheric lobule VI, BA 19 |
|  |  | -38,-56,-36 | -3.379 | 0.01899999 | Left cerebellum, crus I |
|  |  | -14,-66,-24 | -3.375 | 0.00099999 | Left cerebellum, hemispheric lobule VI |
|  |  | 2,-84,-28 | -3.308 | 0.00199997 | Left cerebellum, crus II |
|  |  | 20,-66,-24 | -3.154 | 0.00199997 | Right cerebellum, hemispheric lobule VI, BA 19 |
|  |  | -30,-62,-38 | -3.134 | 0.02200002 | (undefined) |
|  |  | 12,-38,-28 | -3.105 | 0.00199997 | (undefined) |
|  |  | -18,-70,-34 | -3.094 | 0.00199997 | Left cerebellum, crus I |
|  |  | 24,-44,-34 | -3.091 | 0.028 | Middle cerebellar peduncles |
|  |  | 0,-88,-20 | -3.055 | 0.01300001 | (undefined) |
|  |  | 28,-58,-40 | -3.025 | 0.00700003 | Middle cerebellar peduncles |
|  |  | 0,-38,-4 | -3.005 | 0.005 | Cerebellum, vermic lobule III |
|  |  | 2,-24,-8 | -2.993 | 0.00999999 | (undefined) |
|  |  | -2,-92,-18 | -2.951 | 0.01300001 | (undefined), BA 18 |
|  |  | -14,-28,-10 | -2.943 | 0.00999999 | Left hippocampus |
|  |  | -46,-62,-32 | -2.937 | 0.02200002 | Left cerebellum, crus I |
|  |  | -14,-84,-30 | -2.922 | 0.023 | Left cerebellum, crus II |
|  |  | 2,-28,-6 | -2.913 | 0.00999999 | (undefined) |
|  |  | 0,-36,0 | -2.903 | 0.00599998 | (undefined) |
|  |  | -46,-68,-30 | -2.899 | 0.02200002 | Left cerebellum, crus I |
|  |  | -42,-62,-34 | -2.877 | 0.023 | Left cerebellum, crus I |
|  |  | 0,-26,-22 | -2.776 | 0.03100002 | (undefined) |
|  |  | -30,-76,-32 | -2.747 | 0.02399999 | Left cerebellum, crus I |
|  |  | -14,-88,-26 | -2.737 | 0.02899998 | Left cerebellum, crus I |
|  |  | 4,-26,-22 | -2.732 | 0.03200001 | Right cortico-spinal projections |
|  |  | 2,-28,4 | -2.699 | 0.02399999 | (undefined) |
|  |  | -4,-28,-10 | -2.697 | 0.00999999 | (undefined) |
|  |  | -22,-76,-32 | -2.694 | 0.00999999 | Left cerebellum, crus I |
|  |  | 2,-94,-12 | -2.67 | 0.01599997 | (undefined), BA 17 |
|  |  | 4,-20,-2 | -2.659 | 0.00999999 | (undefined) |
|  |  | -26,-76,-32 | -2.641 | 0.02399999 | Left cerebellum, crus I |
|  |  | -6,-80,-14 | -2.58 | 0.01300001 | Left cerebellum, hemispheric lobule VI, BA 17 |
|  |  | -44,-78,-24 | -2.57 | 0.02999997 | Left cerebellum, crus I, BA 19 |
|  |  | -8,-88,-26 | -2.561 | 0.01700002 | Left cerebellum, crus II |
|  |  | -40,-80,-26 | -2.532 | 0.02999997 | Left cerebellum, crus I, BA 19 |
|  |  | 4,-36,4 | -2.5 | 0.028 | (undefined) |
|  |  | -2,-26,-14 | -2.429 | 0.028 | Left cortico-spinal projections |
|  |  | 18,-84,-24 | -2.394 | 0.04500002 | Right cerebellum, crus I, BA 18 |
|  |  | -42,-82,-20 | -2.382 | 0.02999997 | (undefined), BA 19 |
|  |  | 14,-78,-24 | -2.313 | 0.04500002 | Right cerebellum, crus I, BA 18 |
|  | 2 | -2,-60,44 | -3.849 | 0.00999999 | Left precuneus |
|  |  | 6,-56,58 | -3.841 | 0.009 | Right precuneus, BA 5 |
|  |  | 0,-56,50 | -3.815 | 0.00999999 | Left precuneus |
|  |  | 0,-58,54 | -3.705 | 0.00999999 | Left precuneus, BA 7 |
|  |  | 4,-62,56 | -3.526 | 0.00999999 | Right precuneus, BA 7 |
|  |  | 0,-66,42 | -3.504 | 0.00999999 | Left precuneus, BA 7 |
|  |  | 10,-62,44 | -3.43 | 0.00999999 | Corpus callosum |
|  |  | 8,-66,46 | -3.355 | 0.00999999 | Right precuneus, BA 7 |
|  |  | 12,-66,40 | -3.203 | 0.014 | Corpus callosum |
|  |  | -8,-56,52 | -3.176 | 0.01999998 | Left precuneus |
|  |  | -6,-70,40 | -3.094 | 0.028 | Left precuneus, BA 7 |
|  |  | -10,-70,38 | -3.066 | 0.028 | Left precuneus, BA 7 |
|  |  | 4,-74,42 | -3.022 | 0.01700002 | Right precuneus, BA 7 |
|  |  | 8,-58,70 | -3.007 | 0.02999997 | Right precuneus, BA 5 |
|  | 3 | -54,-28,-10 | -4.309 | 0.00999999 | Left arcuate network, long segment |
|  |  | -54,-22,-14 | -3.911 | 0.01300001 | Corpus callosum |
|  |  | -56,-52,10 | -3.739 | 0.01899999 | Left middle temporal gyrus, BA 21 |
|  |  | -54,-42,-8 | -3.574 | 0.02399999 | Left arcuate network, long segment |
|  |  | -66,-48,-8 | -3.064 | 0.03200001 | Left middle temporal gyrus, BA 37 |
|  |  | -46,-42,-12 | -2.862 | 0.04500002 | Left inferior network, inferior longitudinal fasciculus |
|  | 4 | 2,-8,8 | -4.072 | 0.014 | (undefined) |
|  |  | 2,-2,4 | -3.977 | 0.014 | Right anterior thalamic projections |

*Abbreviations: BA* – Brodmann Area, *HC* – healthy controls*, pFWE* –family-wise error corrected p-value, *SDM* – seed-based d-mapping.

Table S5 – Jack-knife sensitivity analyses, heterogeneity and bias in the early psychosis meta-analysis.

| **Contrast** | **Cluster** | **Region** | **Meta-bias test** | ***I^2^*** | **N iterations significant** |
| --- | --- | --- | --- | --- | --- |
| Early psychosis < HC | 1 | Bilateral MSFG, ACC | Bias: 0.01, z: 0.01, df: 3, p: 0.996 | 0.22 | 3/5 (NS(Kyriakopoulos et al., 2012; Li et al., 2019)) |
|  | 2 | R Caudate nucleus | Bias: 0.04, z: 0.01, df: 3, p: 0.990 | 1.75 | 3/5 (NS(Li et al., 2019; Scheuerecker et al., 2008)) |
|  | 3 | L IFG, opercular | Bias: -0.00, z: -0.00, df: 3, p: 0.999 | 1.05 | 2/5 (NS(Kyriakopoulos et al., 2012; Li et al., 2019; Yoo et al., 2005)) |
|  | 4 | L Caudate nucleus | Bias: 0.04, z: 0.01, df: 3, p: 0.991 | 0.63 | 1/5 (NS in all except(Nielsen et al., 2017)) |
|  | 5 | L IFG, triangular | Bias: -0.00, z: -0.00, df: 3, p: 0.999 | 0.16 | 1/5 (NS in all except(Nielsen et al., 2017)) |

*Abbreviations: HC* – healthy controls, *MSFG* – medial superior frontal gyrus, *ACC* – anterior cingulate cortex, *R* – right, *L* – left, *IFG* – inferior frontal gyrus, *MNI* – Montreal Neuroimaging Institute, *BA –* Brodmann Area, *SDM* – seed-based d-mapping, *NS* – when removing the studies cited, the cluster became non-significant (jack-knife analysis).

Table S6 – Jack-knife sensitivity analyses, heterogeneity and bias in the chronic psychosis meta-analysis.

| **Contrast** | **Cluster** | **Region** | **Meta-bias test** | ***I^2^*** | **N iterations significant** |
| --- | --- | --- | --- | --- | --- |
| Chronic psychosis > HC | 1 | Bilateral ACC, MSFG, Insula, Hippocampus | Bias: -0.01, z: -0.02, df: 28, p: 0.986 | 1.57 | 29/30  (NS: (Tik et al., 2021)) |
|  | 2 | Bilateral Median, Posterior cingulate | Bias: 0.02, z: 0.03, df: 28, p: 0.979 | 0.25 | 29/30  (NS: (Tik et al., 2021)) |
|  | 3 | L Superior temporal gyrus | Bias: -0.01, z: -0.02, df: 28, p: 0.988 | 8.83 | 26/30  (NS:(Kaminski et al., 2019; Schlagenhauf et al., 2010; Tik et al., 2021; Wu et al., 2017)) |
|  | 4 | L Supramarginal gyrus | Bias: 0.00, z: 0.00, df: 28, p: 1.000 | 0.02 | 26/30  (NS:(Kaminski et al., 2019; Royer et al., 2009; Schlagenhauf et al., 2010; Tik et al., 2021)) |
| Chronic psychosis < HC | 1 | Cerebellum, thalamus | Bias: 0.00, z: 0.00, df: 28, p: 0.999 | 13.60 | 30/30 |
|  | 2 | Bilateral Precuneus | Bias: 0.01, z: 0.01, df: 28, p: 0.994 | 31.93 | 28/30  (NS:(Guardiola-Ripoll et al., 2022; Pomarol-Clotet et al., 2008)) |
|  | 3 | L Middle temporal gyrus | Bias: 0.00, z: 0.00, df: 28, p: 0.999 | 0.28 | 28/30  (NS:(Pomarol-Clotet et al., 2008; Tik et al., 2021)) |
|  | 4 | Thalamus | Bias: 0.01, z: 0.01, df: 28, p: 0.994 | 0.28 | 26/30  (NS:(Guse et al., 2013; Kaminski et al., 2019; Pomarol-Clotet et al., 2008; Schlagenhauf et al., 2010)) |

*Abbreviations: HC* – healthy controls, *MSFG* – medial superior frontal gyrus, *ACC* – anterior cingulate cortex, *R* – right, *L* – left, *MNI* – Montreal Neuroimaging Institute, *BA –* Brodmann Area, *SDM* – seed-based d-mapping, *NS* – when removing the studies cited, the cluster became non-significant (jack-knife analysis).

Table S7 – Results of the meta-analysis in the at-risk groups with no covariates. Results are shown at uncorrected p-value < .005, cluster size > 10 voxels.

| **Contrast** | **Cluster** | **MNI coordinates** | **SDM-Z** | **p*_uncorr._*** | **Voxels** | **GM region** |
| --- | --- | --- | --- | --- | --- | --- |
| FHR > HC | *No suprathreshold clusters.* | | | | | |
| FHR < HC | 1 | 12,-44,4 | –3.839 | 0.00006187 | 98 | Right lingual gyrus, BA 27 |
| CHR-P > HC | *No suprathreshold clusters.* | | | | | |
| CHR-P < HC | *No suprathreshold clusters.* | | | | | |
| at-risk (FHR+CHR-P) > HC | 1 | 38,52,-4 | 3.528 | 0.00020903 | 111 | Right middle frontal gyrus, orbital part, BA 46 |
|  | 2 | 44,-60,38 | 3.004 | 0.00133121 | 25 | Right angular gyrus, BA 39 |
| at-risk (FHR+CHR-P) < HC | 1 | 14,40,42 | –2.967 | 0.00150222 | 15 | Right superior frontal gyrus, medial, BA 9 |

*Abbreviations: CHR-P –* clinical high-risk for psychosis, *FHR* – familial high-risk for psychosis, *HC –* healthy controls, *MNI* – Montreal Neuroimaging Institute, *GM* – grey matter, *BA* – Brodmann Area, *SDM* – seed-based d-mapping.

Table S8– Results of the meta-analysis in the early and chronic psychosis groups with no covariates. Results are shown at corrected p*FWE* < .005, cluster size > 10 voxels.

| **Contrast** | **Cluster** | **MNI coordinate** | **SDM-Z** | **p*FWE*** | **Voxels** | **GM region** |
| --- | --- | --- | --- | --- | --- | --- |
| Early psychosis < HC | 1 | 2,32,34 | –3.674 | <.001 | 1866 | Left superior frontal gyrus, medial, BA 32 |
|  | 2 | 10,6,12 | –4.224 | 0.008 | 82 | Right caudate nucleu |
| Early psychosis > HC | *No suprathreshold clusters* | | | | | |
| Chronic psychosis < HC | 1 | 0,-54,-26 | – 4.766 | 0.002 | 824 | Cerebellum |
| Chronic psychosis > HC | 1 | 6,32,-8 | 4.926 | <.001 | 3043 | Right anterior cingulate / paracingulate gyri, BA 11 |
|  | 2 | 40,-22,14 | 4.662 | 0.004 | 248 | Right heschl gyrus, BA 48 |

*Abbreviations: HC –* healthy controls, *MNI* – Montreal Neuroimaging Institute, *GM* – grey matter, *BA* – Brodmann Area, *SDM* – seed-based d-mapping, *pFWE* –family-wise error corrected p-value.

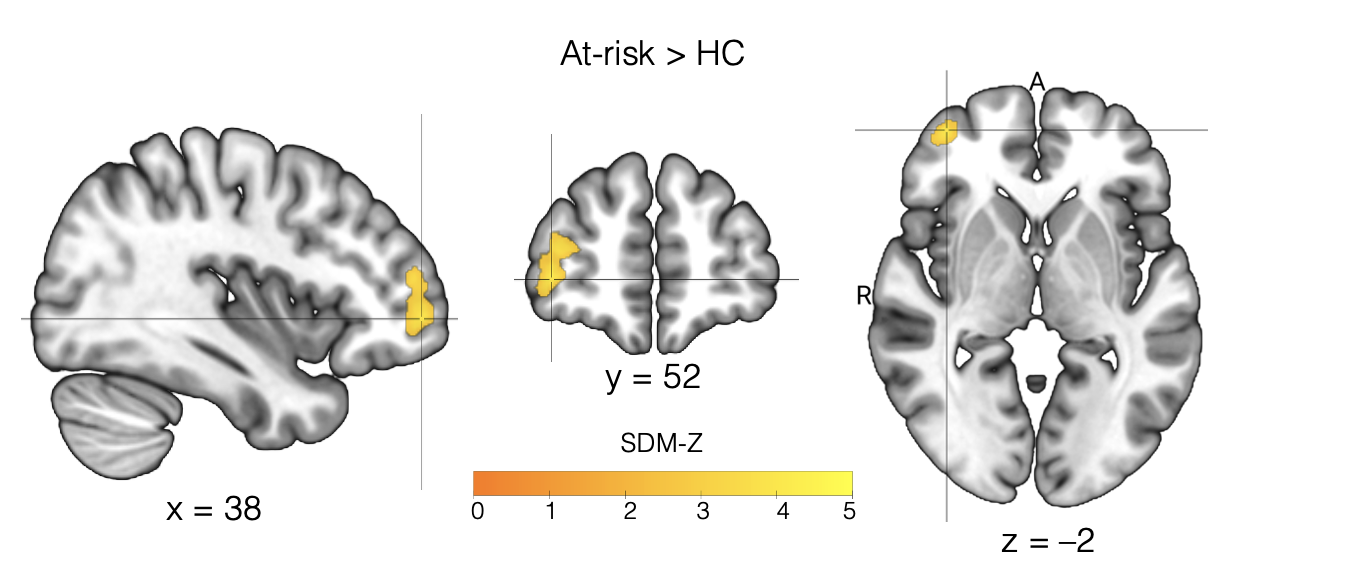
Figure S1 – Meta-analytic results for at-risk for psychosis compared to healthy controls. The cluster emerged at the uncorrected p < .005 threshold, with a cluster extent limit of 100 voxels.

*Abbreviations: A* – anterior, *HC* – healthy controls, *SDM –* seed-based d-mapping, *R* – right.

Figure S2 – Meta-comparison results for at-risk for psychosis compared to early psychosis controls. The cluster emerged at the uncorrected p < .0005 threshold; the same threshold was used for illustrative purposes.

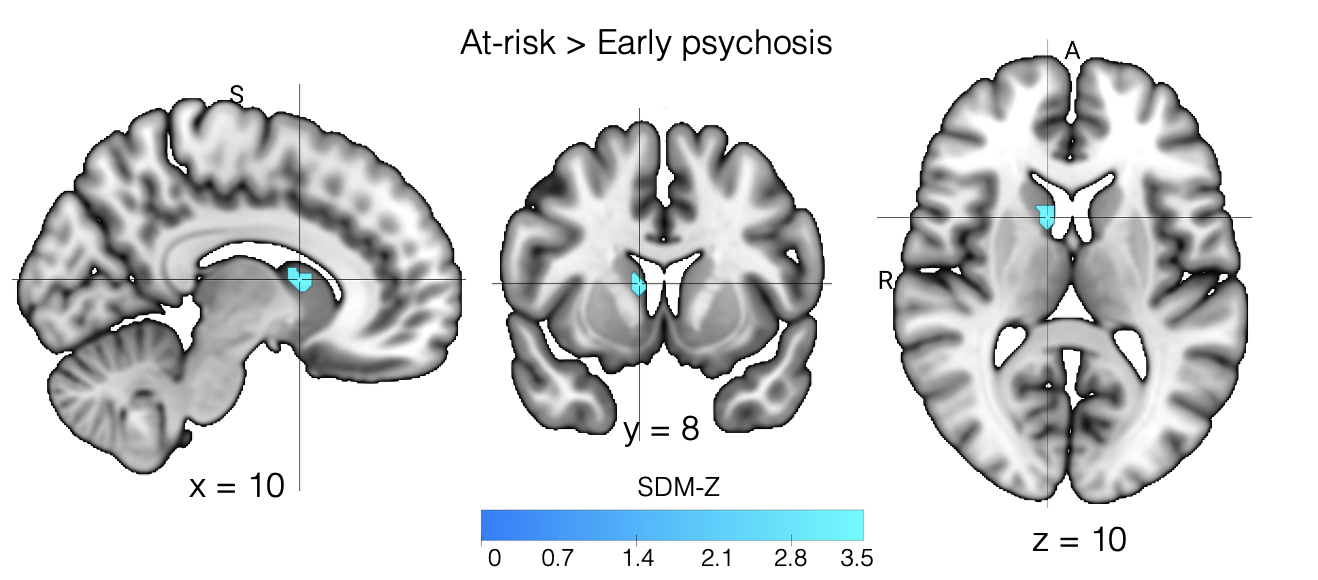

*Abbreviations: SDM –* seed-based d-mapping, *A* – anterior, *R* – right, *S* – superior.

Figure S3 – Meta-regression results effects of sex on brain activity during WM in chronic psychosis (compared to controls). The clusters emerged at the uncorrected p < .0005 threshold; the same threshold was used for illustrative purposes.

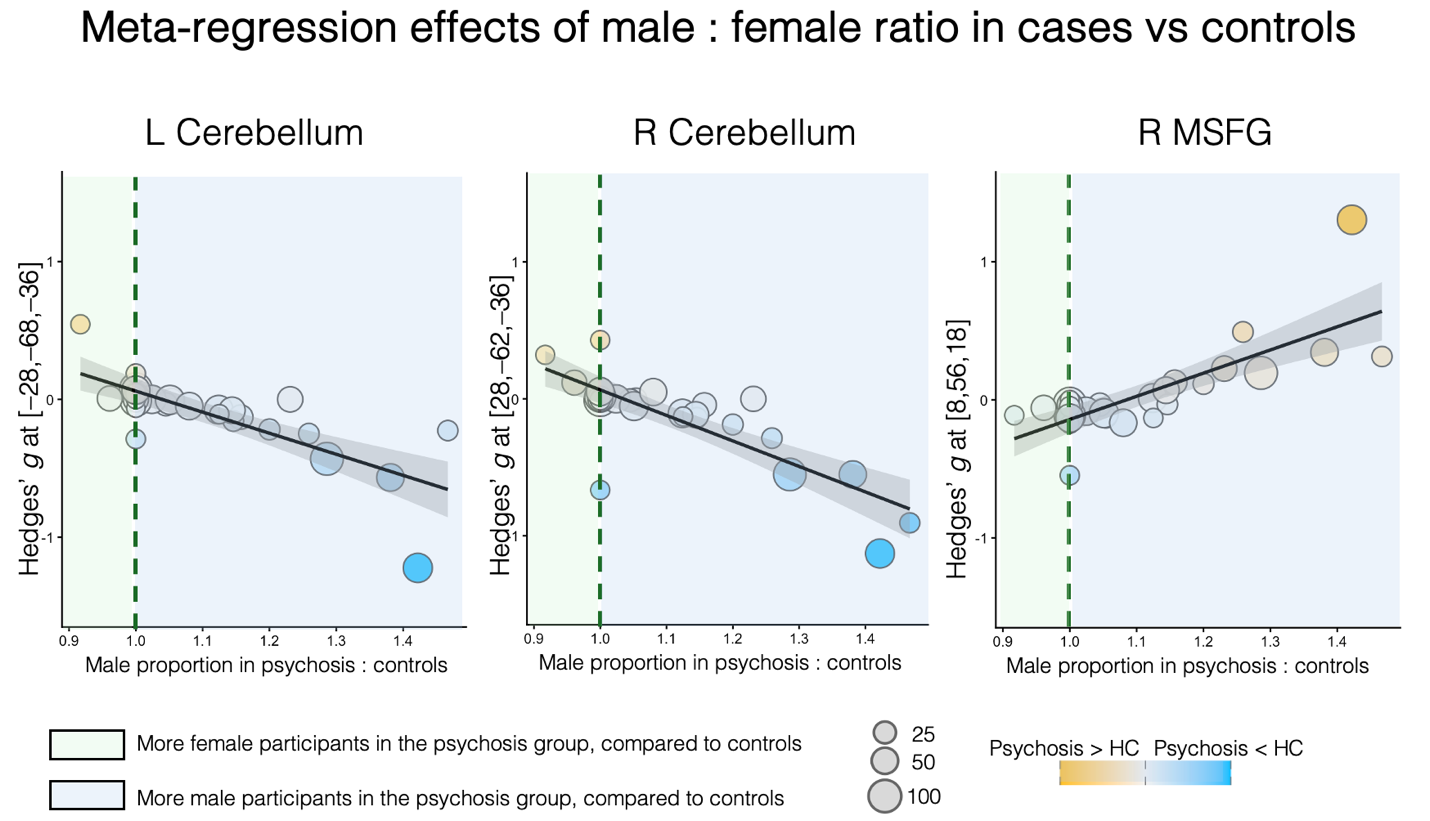

*Abbreviations: HC –* healthy controls, *L* – left, *R* – right, *MSFG* – medial superior frontal gyrus.

van Gool, K.C.A., Collin, G., Bauer, C.C.C., Molokotos, E., Mesholam-Gately, R.I., Thermenos, H.W., Seidman, L.J., Gabrieli, J.D.E., Whitfield-Gabrieli, S., Keshavan, M.S., 2022. Altered working memory-related brain activity in children at familial high risk for psychosis: A preliminary study. Schizophr. Res.

Walter, H., Wunderlich, A.P., Blankenhorn, M., Schafer, S., Tomczak, R., Spitzer, M., Gron, G., 2003. No hypofronatality, but absence of prefrontal lateralization comparing verbal and spatial working memory in schizophrenia. Schizophr. Res.

Wang, X., Cheng, B., Roberts, N., Wang, S., Luo, Y., Tian, F., Yue, S., 2021. Shared and distinct brain fMRI response during performance of working memory tasks in adult patients with schizophrenia and major depressive disorder. Hum. Brain Mapp. 42, 5458–5476. https://doi.org/10.1002/hbm.25618

Williams, J.C., Zheng, Z.J., Tubiolo, P.N., Luceno, J.R., Gil, R.B., Girgis, R.R., Slifstein, M., Abi-Dargham, A., Van Snellenberg, J.X., 2023. Medial Prefrontal Cortex Dysfunction Mediates Working Memory Deficits in Patients With Schizophrenia. Biol. Psychiatry Glob. Open Sci. 3, 990–1002. https://doi.org/10.1016/j.bpsgos.2022.10.003

Wu, D., Jiang, T., 2020. Schizophrenia-related abnormalities in the triple network: a meta-analysis of working memory studies. Brain Imaging Behav. 14, 971–980. https://doi.org/10.1007/s11682-019-00071-1

Wu, S., Wang, Huiling, Chen, C., Zou, J., Huang, H., Li, P., Zhao, Y., Xu, Q., Zhang, L., Wang, Hesheng, Pandit, S., Dahal, S., Chen, J., Zhou, Y., Jiang, T., Wang, G., 2017. Task Performance Modulates Functional Connectivity Involving the Dorsolateral Prefrontal Cortex in Patients with Schizophrenia. Front. Psychol.

Yao, Y., Zhang, S., Wang, B., Lin, X., Zhao, G., Deng, H., Chen, Y., 2024. Neural dysfunction underlying working memory processing at different stages of the illness course in schizophrenia: a comparative meta-analysis. Cereb. Cortex 34, bhae267. https://doi.org/10.1093/cercor/bhae267

Yaple, Z.A., Tolomeo, S., Yu, R., 2021. Mapping working memory-specific dysfunction using a transdiagnostic approach. NeuroImage Clin. 31, 102747. https://doi.org/10.1016/j.nicl.2021.102747

Yoo, S.S., Choi, B.G., Juh, R.H., Park, J.M., Pae, C.U., Kim, J.J., Lee, S.J., Lee, C., Paik, I.H., Lee, C.U., 2005. Working memory processing of facial images in schizophrenia: fMRI investigation. Int J Neurosci.

Zhang, R., Picchioni, M., Allen, P., Toulopoulou, T., 2016. Working Memory in Unaffected Relatives of Patients With Schizophrenia: A Meta-Analysis of Functional Magnetic Resonance Imaging Studies. Schizophr. Bull. 42, 1068–1077. https://doi.org/10.1093/schbul/sbv221
